# Multimodal Clustering Analysis of meta-analytically derived brain regions in Schizophrenia Spectrum Disorders

**DOI:** 10.1101/2025.04.18.25325823

**Authors:** Maxim Korman, Keith M. Smith, Tengjia Jiang, Sergi Papiol, Berkhan Karsli, Genc Hasanaj, Marcel S. Kallweit, Alexandra Hisch, Verena Meisinger, CDP Working Group, Joanna Moussiopoulou, Vladislav Yakimov, Emanuel Boudriot, Michael J. Ziller, Andrew Zalesky, Andrea Schmitt, Peter Falkai, Elias Wagner, Florian Raabe, Lukas Roell, Daniel Keeser

**Affiliations:** Department of Psychiatry and Psychotherapy, LMU University Hospital, LMU Munich; Munich, Germany; Neuroimaging Core Unit (NICUM) University Hospital LMU Munich; Munich, Germany; Department of Computer and Information Sciences, University of Strathclyde; Glasgow, United Kingdom; Max Planck Institute of Psychiatry; Munich Germany; Department of Psychiatry, University of Münster, 48149 Münster, Germany; Center for Soft Nanoscience, University of Münster, 48149 Münster, Germany; Melbourne Neuropsychiatry Centre, Department of Psychiatry, The University of Melbourne, Victoria, Australia; Department of Biomedical Engineering, The University of Melbourne, Victoria, Australia; Laboratory of Neuroscience (LIM27), Institute of Psychiatry, University of Sao Paulo, São Paulo, Brazil; German Center for Mental Health (DZPG), partner site Munich/Augsburg; Evidence-based psychiatry and psychotherapy, Faculty of Medicine, University of Augsburg, Augsburg, Germany; Department of Psychology, Ludwig-Maximilians-University Munich, Munich, Germany; Munich Center for Neurosciences (MCN), Ludwig Maximilian University LMU, Munich, Germany

## Abstract

**Background:** Schizophrenia spectrum disorders (SSD) present substantial clinical and biological heterogeneity, impeding advances in diagnostic precision and personalised treatment. Despite consistent evidence of brain alterations, the identification of subgroups reflecting the disorders’ complexity remains challenging, due to interindividual variability and the inherent limitations of studying neuroimaging modalities in isolation. We developed a novel meta-analytically anchored clustering approach integrating structural (T1-weighted, diffusion tensor imaging) and functional (resting-state fMRI) brain data to derive multimodal subgroups.

**Methods:** We analysed data from 146 SSD patients and 129 healthy controls, initially replicating gray matter volume alterations reported in meta-analyses. We then examined these regions across neuroimaging modalities and performed multimodal clustering on 104 patients with complete data across modalities. The clusters were probed for validity and robustness and characterised by clinical features, polygenic risk scores (PRS) and gene expression pathways.

**Results:** We successfully replicated gray matter volume alterations in 30/33 regions, with approximately half showing significant cross-modal abnormalities. Thalamic dysfunction emerged as particularly prominent. Clustering identified five distinct SSD subgroups with divergent brain-symptom-genetics profiles. Most notably, one subgroup exhibited pronounced white matter decline with aberrant neuroinflammation and myelin gene expression, while another subgroup showed increased gray matter volumes and elevated PRS for intracranial volume, even exceeding levels in our healthy controls.

**Conclusion:** These findings highlight the relevance of analyzing meta-analytically validated regions across multiple imaging modalities. Our clustering approach successfully identified neurobiologically distinct SSD subgroups, offering a promising framework for addressing the challenge of heterogeneity in severe mental illness and advancing precision psychiatry.

## INTRODUCTION

Schizophrenia spectrum disorders (SSD) rank among the most severe mental health conditions, profoundly affecting patients’ quality of life and life expectancy (1), creating substantial challenges for their families (2) and economies worldwide (3,4). While the global prevalence is estimated around 23.2 million (5), a recent study suggests numbers may be severely underestimated even in developed countries, particularly when including underserved populations such as those in prisons or homeless (6).

Patients with SSD experience psychotic symptoms, negative symptoms such as social withdrawal and loneliness, reduced cognitive capacities and a widely varying overall capacity to function in daily life (7,8). Despite the great clinical need, treatment resistance represents a pervasive challenge, leaving many patients with inadequate symptom relief and persistent functional impairments (9).

The high heritability of SSD (10) suggests a significant biological component in its etiology. Genome-wide association studies have provided compelling evidence of the polygenic architecture of SSD. Genetic covariation between schizophrenia and brain phenotypes has been found to be particularly strong in hubs of structural brain networks (11) and early brain development has been suggested to play a mediating role in the genetic risk of schizophrenia (12). Meta-analyses have consistently reported widespread reductions in gray matter volume (GMV) (13–16), diminished white matter integrity (17,18), and altered functional connectivity (19,20) in patients compared with healthy controls.

Yet, these robust group-level findings can obscure substantial interindividual variability. Patients with mental illnesses exhibit considerable heterogeneity across multiple levels, including clinical presentation (21,22), brain structure (23) and function (20). This variability poses significant challenges for patient stratification and the development of targeted interventions.

Among the different methods used to address this heterogeneity, unsupervised clustering is notable for its independence from defined target labels (24), possibly circumventing the problem of suboptimal diagnostic systems and low inter-rater reliability (25). While most neuroimaging studies have clustered based on one modality (26–29), our work builds on emerging evidence that multimodal integration can overcome the limitations of unimodal approaches. For example, the integration of structural and functional connectivity was found to allow for a more comprehensive explanation of different cognitive domains (30–32). Further, multimodal approaches in supervised machine learning enhance predictive accuracy comparable to doubling the sample size (33), a highly relevant aspect given how resource-intensive data collection in neuroimaging is.

Beyond multimodality, the selection of neuroimaging features for clustering should reflect disease-specific rather than sample-specific characteristics. Sampling variability—the random fluctuation of associations between brain and clinical phenotypes across population subsamples—can inflate certain associations (34), potentially leading to findings that fail to generalize. Therefore, recent work suggests using “hybrid analytic methods, which integrate a prior hypothesis (i.e., theory-driven) with data-driven clustering methods”, making use of “theoretical hypotheses to restrict the exploration spaces” (35). To address this challenge, we integrate meta-analytic insights to constrain our exploration space, while also employing nonlinear dimensionality reduction (DR) techniques to capture the inherent complexity of brain data. Despite the well-documented nonlinearity in brain development (36) and function (120), neuroimaging studies have largely relied on linear methods such as Principal Component Analysis, which may inadequately represent cross-modal brain complexity. Nonlinear DR techniques—increasingly common in genetics (37) —have shown to strongly improve clustering accuracy (121) and are central to our approach.

Our pre-registered workflow (Fig. 1) builds on prior work by leveraging meta-analytic insights to constrain the exploration space, which is then examined across multiple neuroimaging modalities and reduced using a novel nonlinear DR technique.

**Fig. 1:**
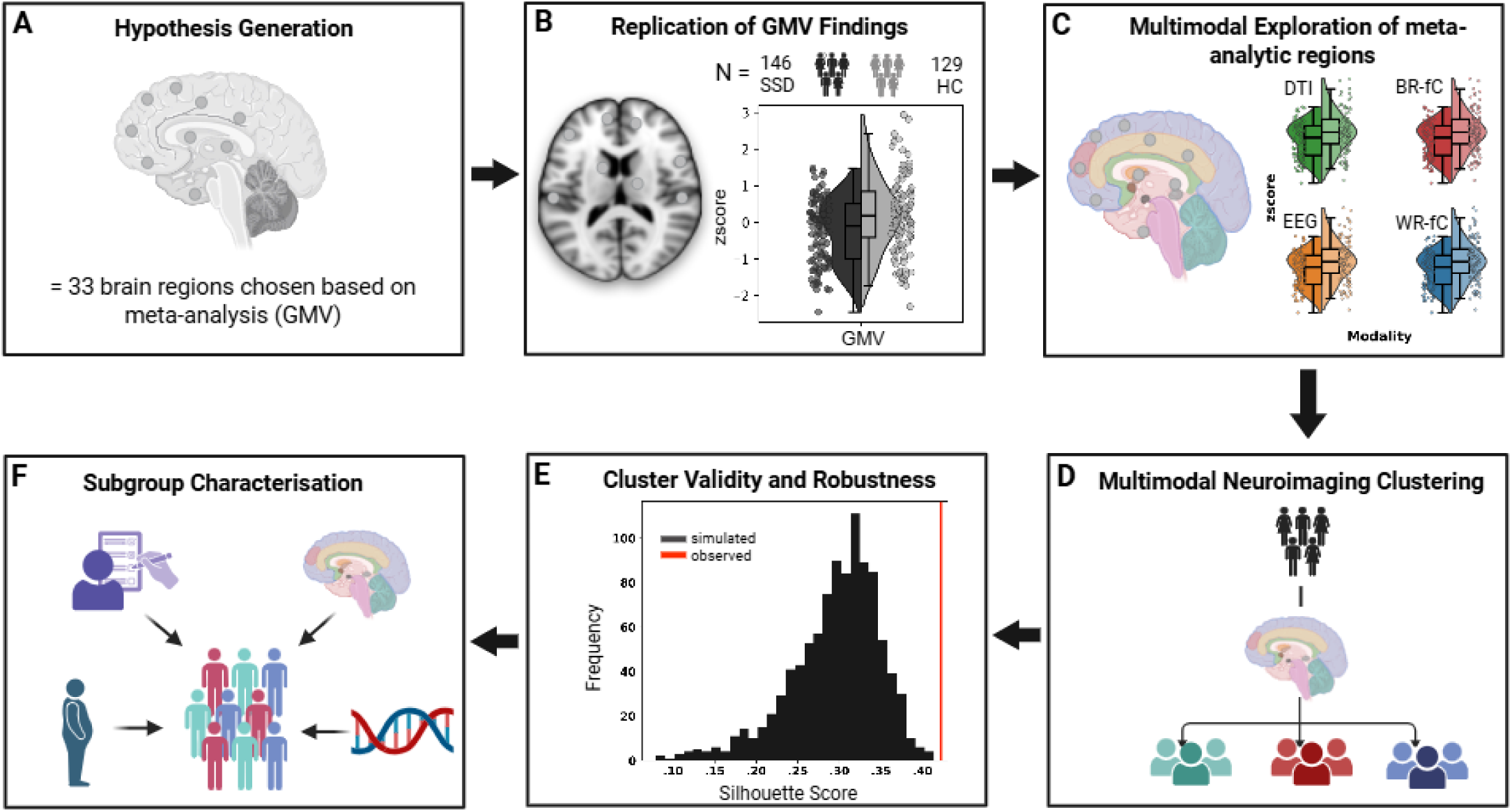
Workflow. **A)** Using pre-registered criteria (https://osf.io/k7wja/), we identified meta-analyses of gray matter changes in SSD, selecting all 33 regions significant in at least two meta-analyses. **B)** These regions were replicated in our cohort and **C)** further explored using a multimodal approach, examining Between-Region functional connectivity (BR-fC), Within-Region functional connectivity (WR-fC), Diffusion Tensor Imaging (DTI), and EEG data. **D)** Pairwise Controlled Manifold Approximation and Projection (PaCMAP) dimensionality reduction and Gaussian Mixture Modeling (GMM) clustering were applied to identify multimodal data-driven subgroups within the patient cohort. **E)** Validity and robustness of the clusters were evaluated through bootstrapping, simulations and additional methods. **E)** Clusters were characterised based on neuroimaging profiles, polygenic risk scores, gene expression pathways, clinical and sociodemographic assessments.

Specifically, we first (i) assess how ROIs identified across meta-analyses of gray matter volume (GMV) replicate in our cohort and (ii) to which extent these regions are impacted across modalities, namely fractional anisotropy (FA), within-region functional connectivity (WR-FC), between-region functional connectivity (BR-FC) and sensor- and source-localized electroencephalography (EEG). Subsequently (iii), we present a novel analytical workflow for multimodal Gaussian Mixture Models (GMM) clustering, aiming to identify robust and clinically meaningful subgroups in a cohort of SSD patients. Lastly, in an exploratory manner, we examine associations of genetic influences and childhood adversity with the derived phenotypes.

To the best of our knowledge, only one previous neuroimaging study has employed a similar nonlinear DR and GMM clustering pipeline, focusing on treatment-resistant depression (38). While that work convincingly demonstrated the potential of this type of analysis, it was limited to structural neuroimaging and included all brain regions as input. Our approach advances this methodology in three key ways: 1) integrating both structural and functional neuroimaging data, 2) implementing a nonlinear DR technique with superior performance in comparative studies (39,40) and 3) specifically targeting meta-analytically derived ROIs in SSD to constrain the feature space, increasing the likelihood of clustering disease-specific rather than sample-specific characteristics.

## Materials and Methods

### Study sample and clinical assessment

This project was conducted as part of the Clinical Deep Phenotyping study (41), an extension of the Mental Health Biobank (ethics project No. 18-716) (42), which received approval from the ethics committee of the Faculty of Medicine, Ludwig Maximilian University Munich (project Nos. 20–0528 and 22-0035) and registered at the German Clinical Trials Register (DRKS00024177). The study included patients diagnosed with schizophrenia (SZ), schizoaffective disorder (SZA), or brief psychotic disorder, as well as healthy controls (HCs) with no lifetime history of psychiatric disorders, as confirmed by the Mini-International Neuropsychiatric Interview (43).

Psychotic symptom severity was evaluated using the Positive and Negative Syndrome Scale (PANSS) (44), while cognitive performance was measured with the Brief Assessment of Cognition in Schizophrenia (BACS) (45). Overall functioning was assessed using the Global Assessment of Functioning (GAF) score (46). In addition, participants completed the Childhood Trauma Screener (CTS) (47), the three-item loneliness scale (66), and the Brief Resilience Scale (BRS) (49). Current antipsychotic medications were converted to chlorpromazine equivalent doses using the defined daily doses method (50).

### ROI selection criteria

In line with our pre-registration, we applied strict inclusion criteria to ensure the reliability of our selected brain regions: We searched pubmed and included only meta-analyses published between 2015 and 2024, using a case-control design with a minimum of 1,500 patients with schizophrenia-spectrum disorders, adults only, whole-brain only, with results for GMV reported in MNI space.

We included all regions that were reported as significant in at least two of the meta-analyses identified, accommodating for differences in study populations and methodologies while focusing on regions with the strongest overall support. To achieve multimodal integration, we employed the Brainnetome (BN) atlas, an atlas that includes volumetric gray matter parcellations, resting-state fMRI (rs-fMRI), and diffusion tensor imaging (DTI). BN regions were then assigned to the meta-analytic regions of interest (ROIs) and at least two meta-analytic MNIs must overlap with a BN region for this region to be included as a ROI. The assignments are shown in Fig 2.

**Fig. 2:**
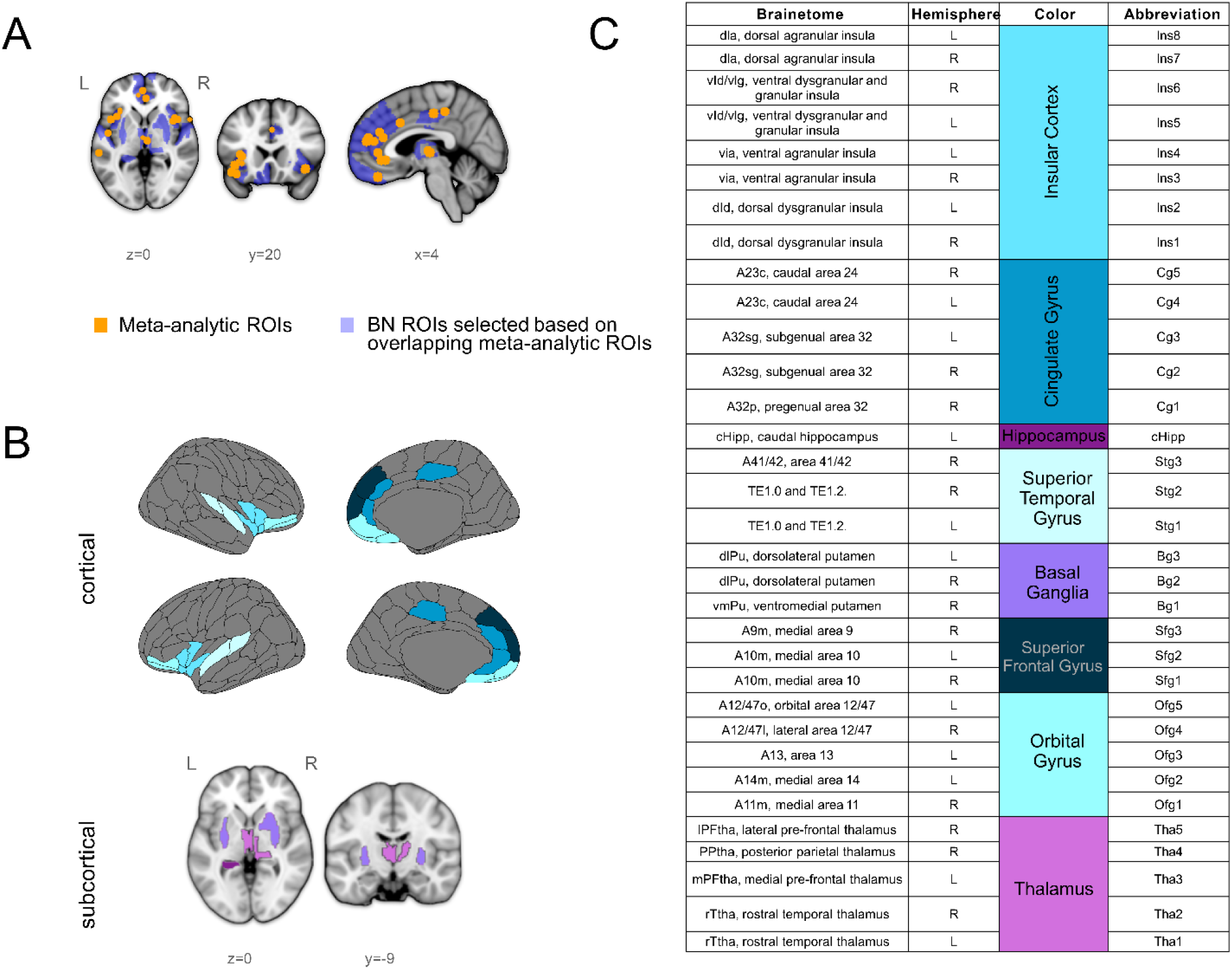
**A)** Meta-analytic ROIs are marked in orange, underlaid with Brainnetome (BN) regions in blue, for all BN areas with at least two corresponding meta-analytic ROIs. **B)** Visualization of our BN ROIs shown for cortical (blue shades) and subcortical areas (violet shades). **C)** All our brain regions listed with abbreviations, corresponding BN names and full names.

### Magnetic Resonance Imaging (MRI)

Clinical scores, neuropsychological and structural T1w and T2w MRI data were shared with the Open Science initiative Psy-ShareD and are publicly available there (https://psyshared.com/). The resting state and DTI data are being prepared for sharing with ENIGMA (https://enigma.ini.usc.edu/). The scanning protocol of the CDP study was based on the HCP Ageing sequences. T1-weighted images, functional EPI images, and DWI images were quality-controlled using the automated quality control software MRIQC (51) combined with visual inspection of all raw images. The T1-weighted images were processed using the Neuromodulation and Multimodal NeuroImaging software (NAMNIs) version 0.3 (52). After standard preprocessing, NAMNIs computes GMVs of brain regions defined by a custom atlas. As outlined previously, we selected the GMVs of meta-analytically derived regions defined by the BN atlas.

Functional resting-state EPI images were preprocessed using fMRIPrep version 23.0.1 (53). Preprocessed images were smoothed (FWHM = 6mm) and denoised using global, cerebrospinal fluid, and white matter signals, along with ICA-AROMA noise components as regressors in Nilearn’s *clean_img* function. Denoising also included band-pass filtering (0.01 – 0.1 Hz), detrending, and standardization. Subject-specific denoised BOLD mean time series were extracted for meta-analytically derived regions of the BN atlas and all voxels within them using Nilearn’s *maskers* module. Functional connectivity was computed in two ways: (1) between region-wise mean time series, yielding a 31 × 31 matrix, averaged column-wise (excluding self-connections) to derive an overall between-region connectivity score (BR-FC); (2) within each region, averaging the upper triangle of the voxel-wise connectivity matrix to quantify intra-region functional connectivity (WR-FC). BR-FC and WR-FC were calculated to cover both global and regional FC, respectively.

DTI data were processed using FMRIB Software Library (FSL) version 6.0.7.8 in combination with tract-based spatial statistics (TBSS) tools. Each subject’s voxel-wise FA map was projected onto the skeleton. The meta-analytically derived regions from the BN atlas were mapped into the individual skeleton and mean FA in each of these regions was calculated.

Finally, GMVs, BR-FC, WR-FC, and mean FA of 33 meta-analytically derived brain regions served as features for the subsequent clustering analysis.

We provide detailed information regarding all preprocessing steps in the supplementary information.

### Electroencephalography

Resting-state EEG was recorded with 32 electrodes using a BrainAmp amplifier (Brain Products, Germany) at 1000 Hz sampling rate, referenced to Cz, and with impedance kept below 5 kΩ. Electrodes followed the International 10/20 system. The 10-minute recording included 5 minutes with eyes closed and 5 minutes with eyes open.

EEG data were preprocessed using a modified pipeline from Adams et al. (54) using EEGLAB v2022.0 (55) (https://sccn.ucsd.edu/eeglab/), separately for eyes-open and eyes-closed conditions.

Source localization was conducted in Brainstorm (Version: April-2024) (56) (https://neuroimage.usc.edu/brainstorm) after EEG preprocessing. Anatomical data processed with FreeSurfer v7.3.2 (57,58) (http://surfer.nmr.mgh.harvard.edu/) were imported into Brainstorm, with the cortical surface set to 15,000 vertices for each subject.

Source estimation used the Linearly Constrained Minimum Variance (LCMV) beamformer (59) with unconstrained dipole orientations, extracting time-series data from 17 cortical BN ROIs, excluding dlPu ROIs, which failed to be reliably imported as scouts for over 40% of subjects.

Power Spectrum Density (PSD) was calculated for time-series of each ROI using the Welch method with 6-second windows and 3-second overlap in MNE-Python (v1.8.0) (60).

### Genotyping, Polygenic Risk Score (PRS) and Pathway Score Calculation

Individuals included in this study were genotyped using Illumina’s Global Screening Array version 3.0 (Life & Brain GmbH). Details on data processing, quality control, and imputation in this cohort can be found elsewhere (61).

Following genotype imputation, genotype dosage data was used to calculate polygenic load for each participant based on the results of the following Genome-wide association studies (GWAS): schizophrenia (62), bipolar disorder (63), educational attainment, volumetric analyses of subcortical areas (putamen, hippocampus, thalamus) and ICV (64). Posterior single nucleotide polymorphism effect sizes were inferred under continuous shrinkage priors using PRS-CS (65), with the global shrinkage parameter (phi) estimated through a fully Bayesian approach (*auto* mode). PLINK v1.90b6.16 was used for PRS calculation using the *--score* function (66).

After genotype imputation, post-imputation quality control was performed to remove SNPs using the following PLINK2 (v2.00a5.12LM) parameters: --maf 0.01, --hwe 1e-6, --geno 0.02. Multiallelic positions were also removed. To harmonize the genotype data with the CASTom-iGEx reference panel, only SNPs present in the reference panel were retained. Dosage data were exported using --export vcf bgz vcf-dosage=DS-only to generate input files (--genoDat_file) for the CASTom-iGEx pipeline (67).

ThPriLer_predictGeneExp_smallerVariantSet_run.R script was used to obtain the predicted gene expression matrix. We used the CommonMind Consortium (CMC) Release (68) composed of RNA-Seq data from post-mortem dorsolateral prefrontal cortex (DLPC) tissue from SSD patients and controls. Tscore_PathScore_diff_run.R was utilized to compute T-scores and pathway scores. Pathway scores were calculated for neuroinflammation, myelin and oxidative pathways (69), for GO pathways 0006979 and 0022010 relating to oxidative stress response and central nervous system myelination, Wikipathways WP4304 and WP2267 relating to oligodendrocyte and glial cell differentiation. These pathways were chosen to examine possible genetic associations with reduced Fractional Anisotropy (FA) in the brain.

### Statistical Analysis and Clustering approach

We applied Ordinary Least Squares (OLS) regression to investigate group differences in the selected brain regions for each region of interest (ROI), comparing patients to controls while including sex as a covariate. For each of the 33 ROIs, we calculated t-statistics and p-values to assess the significance of group differences. False Discovery Rate (FDR) correction was applied across the 33 regressions (one per ROI) to account for multiple comparisons. This process was repeated for each neuroimaging modality.

Regions with FDR-corrected p-values below 0.05 were considered significant. To quantify the magnitude of group differences, we calculated Cohen’s d for each region, representing the standardized effect size between controls and patients.

To analyze if there are multimodal subgroups in our patient population, we applied Pairwise Controlled Manifold Approximation and Projection (PaCMAP) (54) on our multimodal neuroimaging outcomes, followed by clustering with Gaussian Mixture Models (GMM). PaCMAP is a nonlinear DR technique that builds upon methods like t-SNE and UMAP (62), excelling in preservation of local and global structures. We used the default parameters described here (https://github.com/YingfanWang/PaCMAP?tab=readme-ov-file#parameters). Before applying PaCMAP, we used OLS regression to remove the influence of age on brain outcomes, ensuring that age-related variance did not confound the clustering results.

Given that brain outcomes across different modalities and regions vary in range, we z-standardized the residuals to prevent any single variable from disproportionately influencing the clustering process. To perform GMM clustering the sklearn GaussianMixture package was used, using default parameters and the random state set at 42.

To select the optimal number of clusters, we evaluated the quality of different clustering solutions using four validation metrics: Silhouette Score, Calinski-Harabasz Index, Davies-Bouldin Index, and Akaike Information Criterion (AIC). Previous studies have shown that using a combination of 3–5 validation metrics outperforms the worst-performing individual metric in the combination, thereby reducing the risk of misidentifying clusters (71). The cluster solution with the best rank sum of the cluster validation metrics was selected. Further details regarding the validation and robustness of the clusters are found in the supplementary material.

## RESULTS

### Cohort Characteristics

The study sample consisted of 146 patients with a SSD diagnosis (26% female, age = 37.99 ± 11.59) and 129 HCs (51% female, age = 35.85 ± 11.80) (Table 1). The most common diagnosis was schizophrenia (65%), followed by schizoaffective disorder (26%). Patients performed significantly worse than HCs across all clinical measures. For example, BACS scores were lower in patients compared to HCs (SSD = -0.52 ± 0.97 vs HCs = 0.57 ± 0.67, p < 0.001), and patients reported greater loneliness (Loneliness Scale SSD = 7.23 ± 2.31 vs HCs = 5.07 ± 1.43, p < 0.001).

**Table 1:**
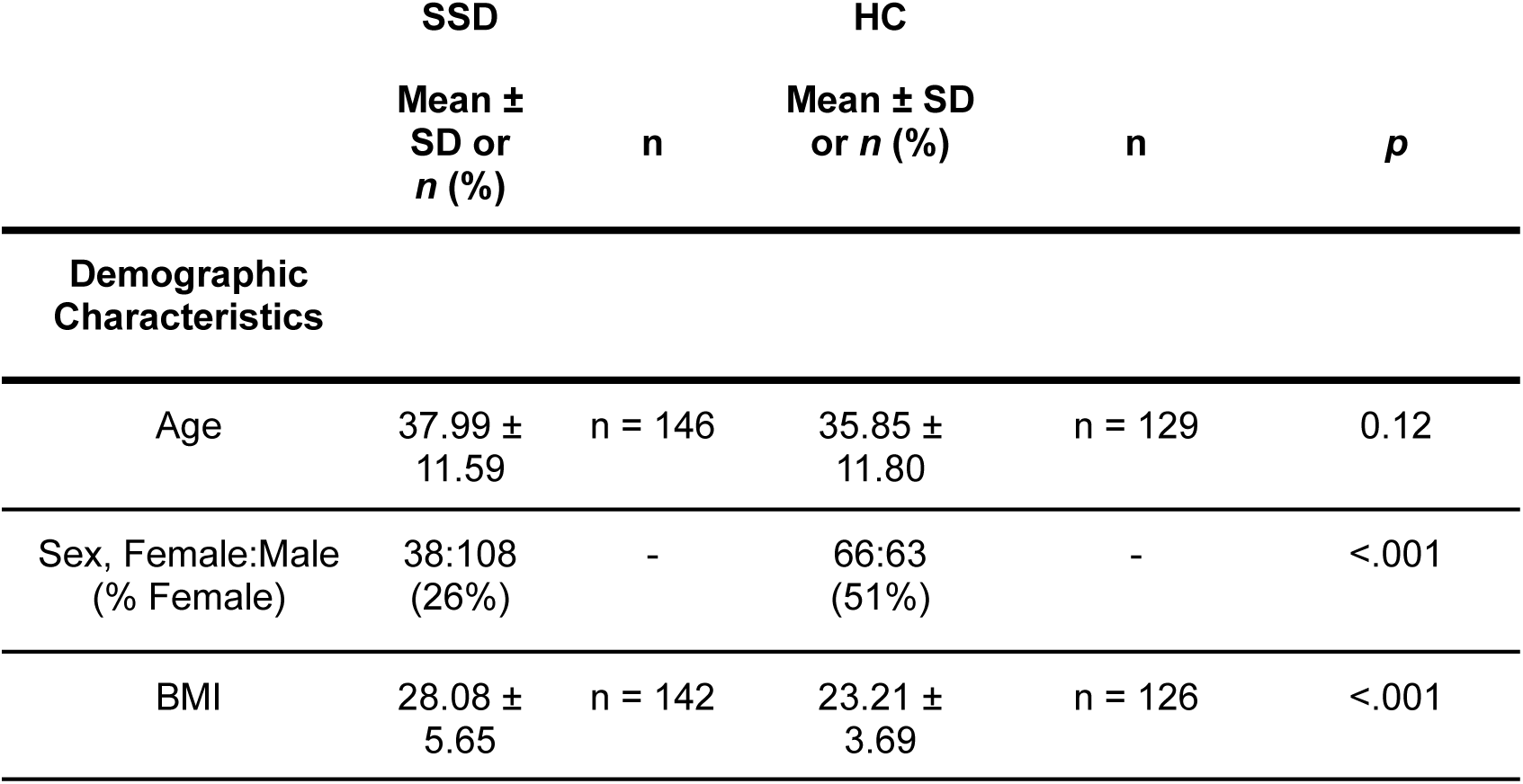
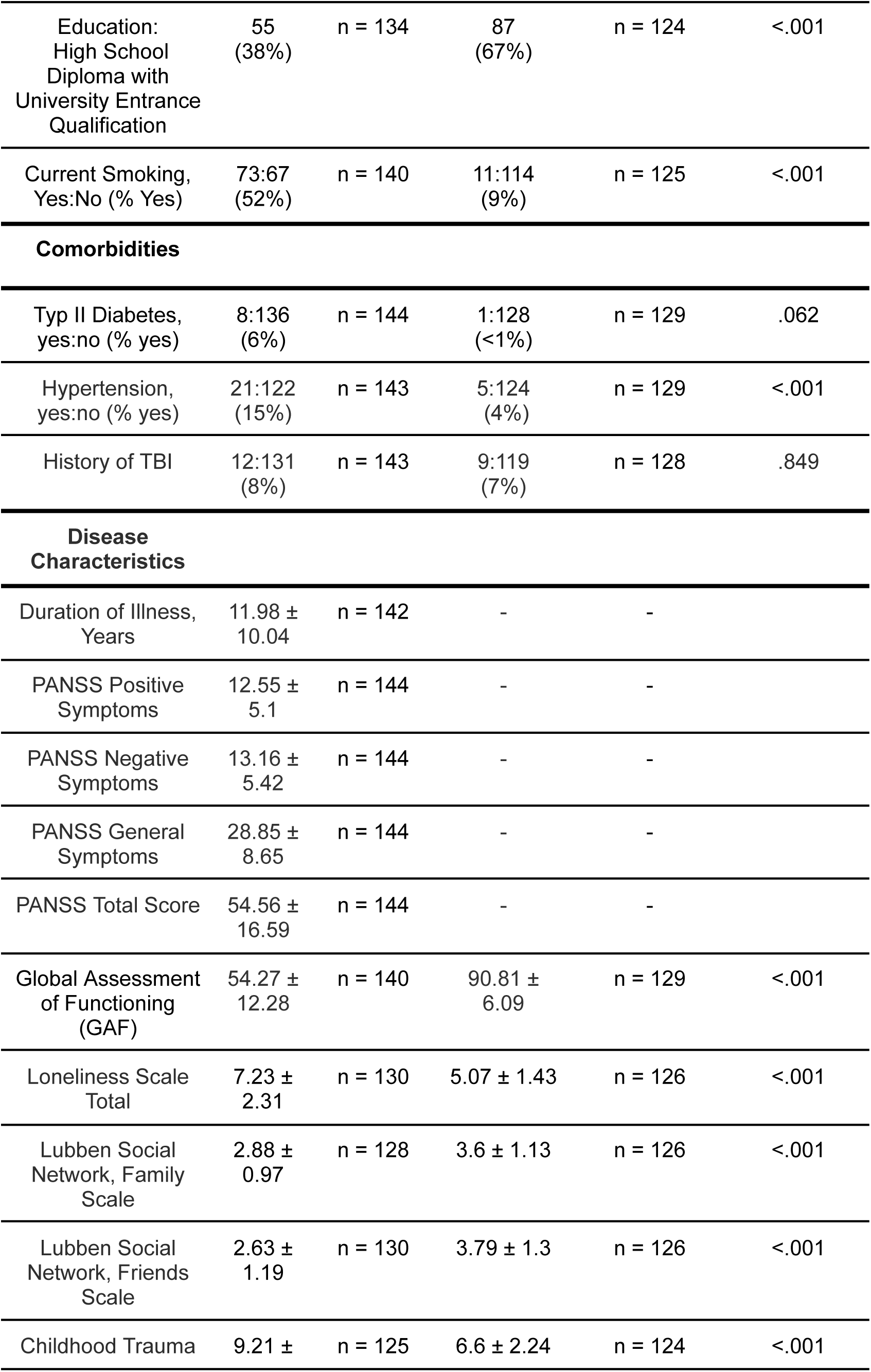
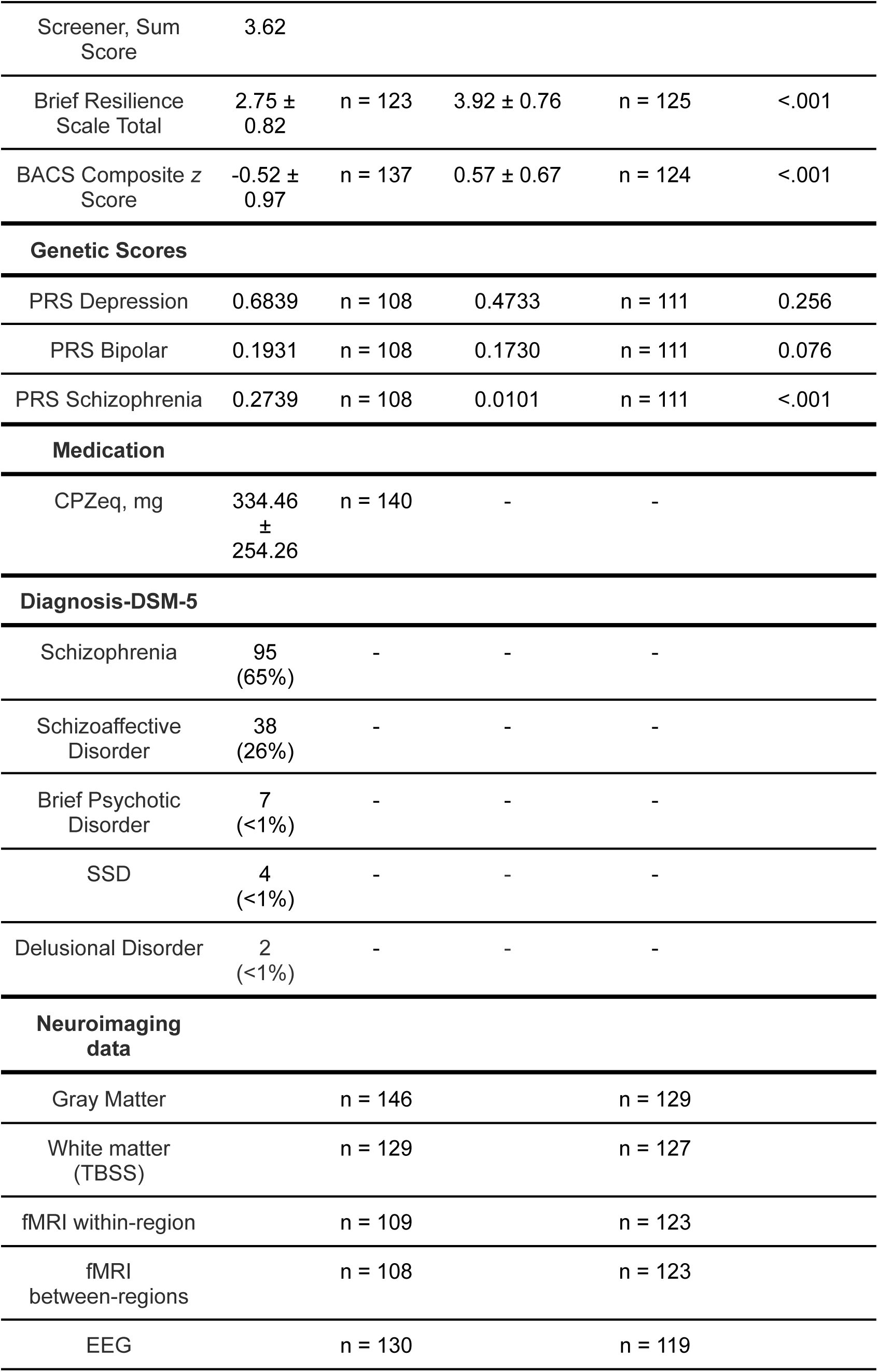
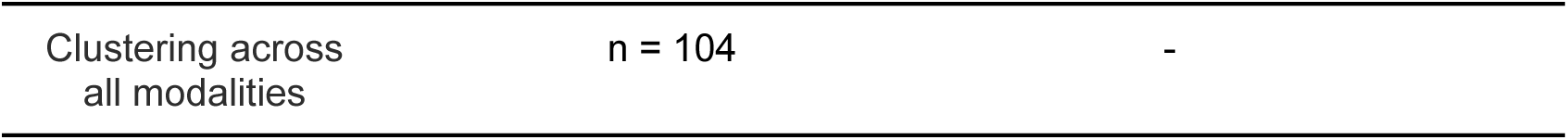
Cohort Characteristics:

Compared to the average SSD patient in large studies (74), patients in this study had relatively high Global Assessment of Functioning (GAF) scores (54.27 ± 12.28) and relatively low Positive and Negative Syndrome Scale (PANSS) scores (54.56 ± 16.59), suggesting a sample with milder symptoms and higher ability to function in daily life.

Both patients and HCs also exhibited high educational achievement compared to the general population, with 38% of patients and 67% of HCs completing high school with an Abitur (university-qualifying diploma), compared to 33.5% in the general population (75).

### Meta-Analyses and selection of ROIs

Applying our search strategy described in the methods section we found six meta-analyses that fulfilled our criteria (13–16,76,77). We selected regions that were shown to significantly differ in their GMV between patients and controls in more than one meta-analysis. These regions include areas from the cingulate cortex (e.g., A32pR, A32sgR, A32sgL), and the insular cortex (e.g., vIaR, vIaL, dIdR, dIdL). Key subcortical regions such as the thalamus (e.g., rTthaL, rTthaR, mPFthaL) and the ventral striatum (e.g., vmPuR, dlPuR) are also represented.

Additionally, regions from the prefrontal cortex (e.g., A10mR, A10mL, A14mL, A9mR), from the orbitofrontal gyrus (A1247lR, A1247oL) and one hippocampal region (cHippL) are among the included regions. A visualization of our ROIs is provided in Fig. 2 and Fig. S1, with a summary table of the meta-analyses provided in the supplementary Table S1.

### Replication of GMV Reductions in Selected Regions

From the 33 ROIs selected based on meta-analyses, 30 regions were found to be significant after applying FDR correction, indicating lower GMV in SSD patients compared to HCs and yielding a very high replication rate of 91%. The ten regions with the highest effect sizes (Cohen’s d) ranged from 0.30 to 0.46. Notably, three regions—the bilateral rostral temporal thalamus and the pregenual area 32 in the cingulate gyrus—had effect sizes greater than 0.4 (d = 0.45 to 0.46), indicating medium to strong effects. A complete list of p-values and effect sizes for all regions is available in table S2.

We conducted a random selection analysis to examine the robustness of our results. For 1000 iterations, we randomly selected 33 regions from the Brainnetome Atlas. The mean number of significant regions across these trials was 13.64 (41%), with no single run yielding more than 25 significant results (Fig. 3B1). Regions chosen based on meta-analyses outperformed random selection by a factor of 2.2.

**Figure 3:**
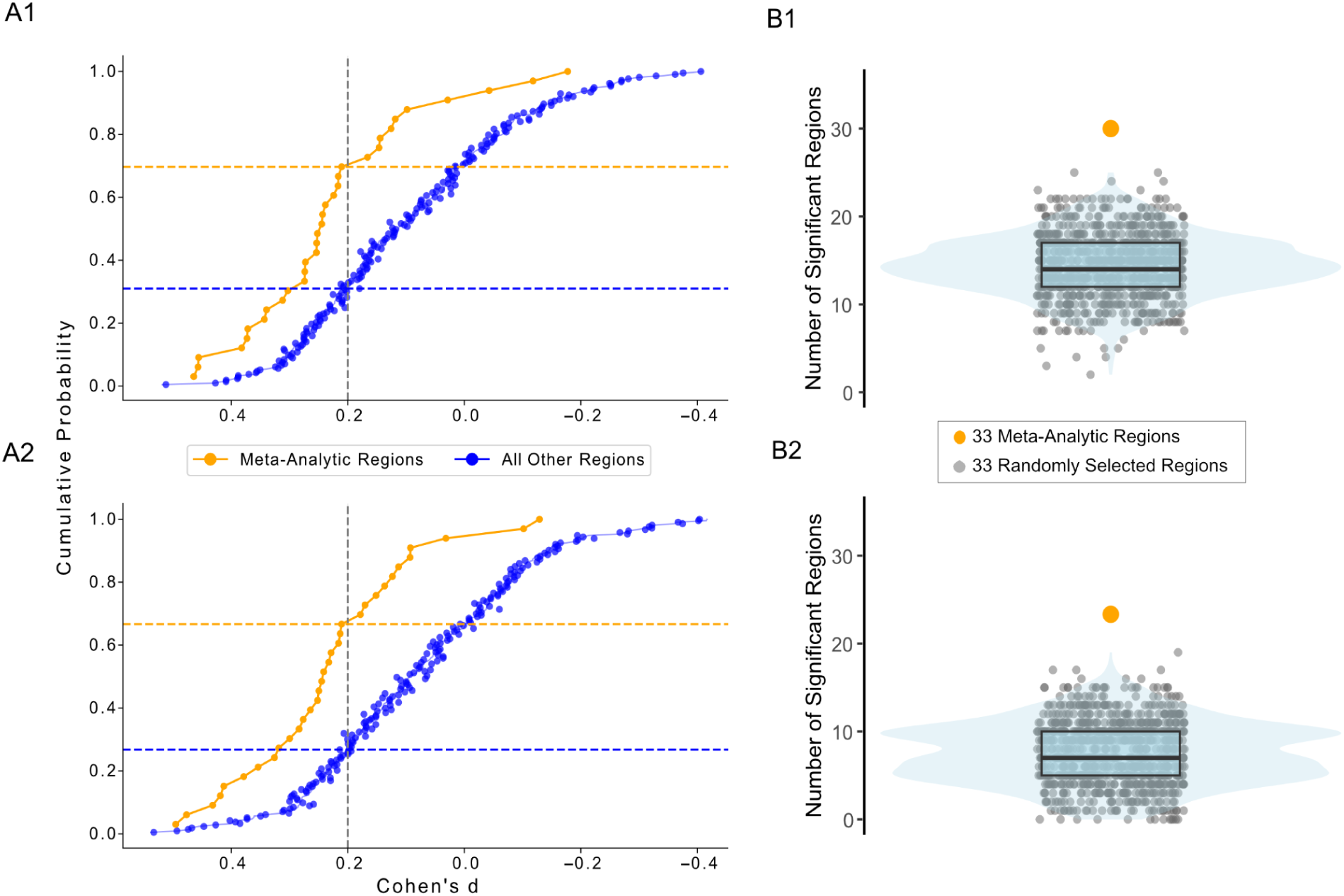
Meta-analysis based ROIs vs. other regions in the Brainnetome Atlas. **Top**: Internal cohort. **Bottom**: Our patients compared with age and sex matched Human Connectome Project Controls. **Left**: Effect sizes of 33 meta-analysis based ROIs vs all regions in the Brainnetome atlas in the internal (a) and external (b) comparison. Cumulative Probability was assessed as the percentage of regions meeting the threshold of a weak effect size (Cohen’s d = 0.2). **Right**: Number of statistically significant regions of 33 meta-analysis based ROIs vs 33 randomly selected regions across 1000 iterations in the internal (a) and external comparison (b).

Furthermore, 70% of meta-analytically derived ROIs, but only 30% of all regions, reach a Cohen’s d of 0.2, defined as the lower end of a small effect size (78) (Fig. 3A1).

We then repeated the same analysis in an external comparison, consisting of 444 exactly age and sex matched HCs (26% female, age = 38.37 ± 11.59), drawn from the Human Connectome Project (HCP Old n = 215; HCP Young n = 178) and the CDP cohort (n = 51). Details regarding site harmonization are provided in the supplementary material. In the external comparison, 22 out of 33 regions showed significant group differences after FDR correction (Table S3). Consistent with our internal cohort, no regions were identified where patients had significantly higher GMV than controls and nine regions exhibited moderate effect sizes (Cohen d = 0.3 - 0.5). Of these nine regions, five had ranked among the highest effect size regions in our internal dataset, reinforcing the robustness of these findings. The most pronounced change observed in the external comparison was in the ventral granular insular gyrus, where effect sizes more than doubled bilaterally compared to the internal cohort.

Random selection in the external comparison produced a mean number of significant regions of 7.7 (23%) (Fig. 3B2). Thus, regions chosen based on meta-analyses outperformed random selection by a factor of 2.9. We found that 67% of meta-analytically derived ROIs and 27% of all other brain regions reached a Cohen’s d of 0.2 (Fig. 3A2), a pattern closely mirroring the internal cohort distribution (70% vs. 30%).

### Case-control Differences Across Imaging Modalities

In the multimodal case-control comparison of our ROIs, 32 out of 33 regions showed significant reductions in fractional anisotropy (FA), 27 regions showed significant reductions in within-region functional connectivity, and 23 regions exhibited significant reductions in between-region functional connectivity, all after FDR correction. In the source-localized EEG analysis of the 17 cortical ROIs, only the right medial area 10 in the superior frontal gyrus showed significant differences between patients and healthy controls during the eyes-closed condition in the low-frequency spectrum (1–12 Hz), with patients exhibiting a higher amplitude. A detailed report of the results for each modality is provided in the supplementary material (S4-S6), and a heatmap of all significant results and effect sizes is presented in Fig.4.

**Figure 4:**
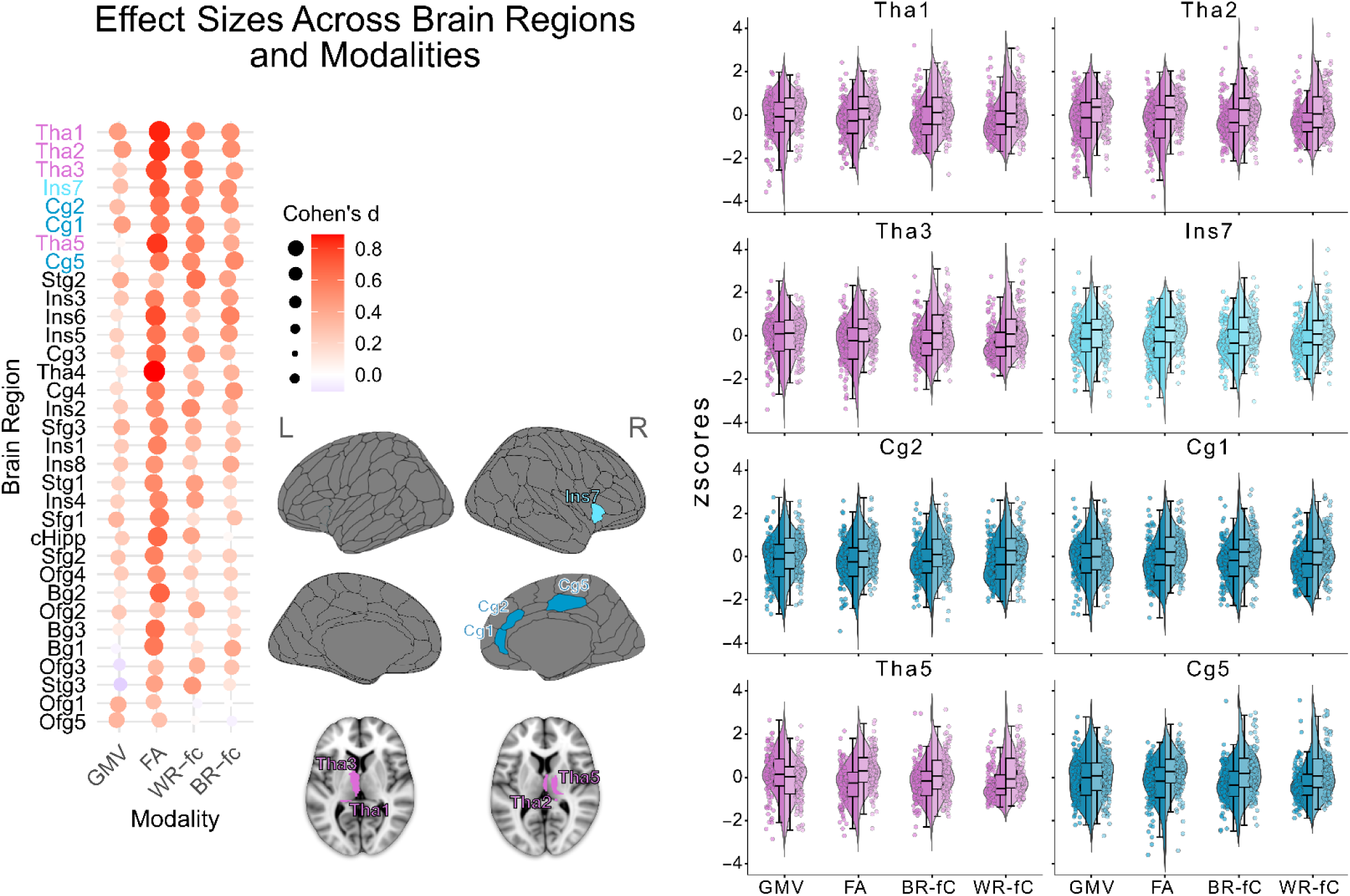
Effect sizes across regions/modalities, Controls vs. Patients. The heatmap left shows all regions across all modalities, sorted by average effect size. The top 8 regions are visualised in the middle and the distribution of outcomes across modalities is depicted on the right. In each violin plot patients are depicted on the left and in darker color. GMV = Gray Matter Volumes; FA = Fractional Anisotropy; WR-fC = Within-Region functional connectivity; BR-fC = Between-Region functional connectivity. Tha1=Left rostral temporal thalamus. Tha2=Right rostral temporal thalamus. Tha3=Left medial pre-frontal thalamus. Ins7=Right dorsal agranular insula. Cg2=Right subgenual area 32. Cg1=Right pregenual area 32. Tha5=Right lateral pre-frontal thalamus. Cg5=Right caudal area 24.

Overall, of the 33 regions selected based on meta-analytic case-control comparisons of GMV, 16 regions showed significant reductions in SSD compared to HCs across four modalities (GMV, FA, WR-fC, BR-fC). Mean effect sizes were highest in thalamic regions, particularly in the bilateral rostral temporal thalamus, which showed consistently high effect sizes across all modalities, with values reaching up to 0.89 in FA. Regions within the cingulate gyrus, notably the pregenual area 32, and the right dorsal agranular insula also showed high mean effect sizes across modalities, though to a lesser extent than the thalamus.

In contrast, regions within the orbital gyrus—specifically, the left orbital area 12/47, medial area 11, and left area 13—demonstrated consistently low effect sizes across all modalities. Medial area 11 was the only region significant in just a single modality.

### Polygenic risk scores and childhood adversity: Case-control difference

Patients exhibited a significantly increased genetic burden for schizophrenia compared to healthy controls (FDR*_p_*= 0.0034), while the PRS for bipolar disorder, major depression and educational attainment were nonsignificant. Among regional volume PRS scores, only the PRS for thalamic volume, higher in HCs, showed a trend toward significance but did not survive FDR correction (FDR*_p_* = 0.099).

Correlation analyses conducted separately for patients and controls revealed that the PRS for thalamic volume had the strongest associations with neuroimaging outcomes. The highest correlations were observed with FA in the left medial prefrontal thalamus (r = 0.25, FDR*_p_* = 0.134) and GMV in the bilateral rostral thalamus (r = 0.24-0.25, FDR*_p_* = 0.134). Interestingly, these correlations were notably higher in HCs compared to patients, suggesting a potential divergence in genetic and structural relationships. Overall, the suggestive differences in the PRS for thalamic volume observed in the case-control comparison, along with its trending correlations with structural brain values in this region, aligns with our findings of very high multimodal thalamic effect sizes. Furthermore, patients had significantly higher CTS scores (SSD = 9.21 ± 3.62, HCs = 6.6 ± 2.24, FDR*_p_* = <.001). The largest differences were observed in emotional abuse (SSD = 2.04 ± 1.22, HCs = 1.31 ± 0.75, FDR*_p_* = <.001) and emotional neglect (SSD = 2.04 ± 1.22, HCs = 1.31 ± 0.75, FDRp = <.001).

### Identification of multimodal clusters in patients with SSD

The clustering analysis was conducted exclusively on patients. Given that the EEG did not contribute to cluster differentiation and its removal increased our sample size from 92 to 104 patients (13%), we decided to proceed without EEG data. Further details on this decision are provided in the supplementary material.

Applying Pairwise Controlled Manifold Approximation (PaCMAP), a nonlinear dimensionality reduction method, on our 133 neuroimaging outcomes (33 regions across 4 modalities) resulted in two projections and a 2-dimensional space showing modality specific patterns. The first PaCMAP projection was positively correlated primarily with GMV, along with some BR-fC outcomes. In contrast, the second PaCMAP projection showed high negative correlations predominantly with FA values, along with a few positive correlations with BR-fC. The pregenual area 32 of the Cingulate Gyrus, the ventromedial putamen, and various insular regions, including the bilateral ventral granular and dysgranular insula, were among the top 10 features most strongly correlated with both PaCMAP projections, contributing significantly to the variance observed in our cohort. Visualizations of this multimodal 2-dimensional projection are provided in Fig. S4.

To identify possible clusters within this space, we used the two PaCMAP projections as input for Gaussian Mixture Models (GMMs). We evaluated models with a range of 1 to 6 clusters, an upper limit chosen with respect to our sample size of 104 patients. The number of clusters were determined through the rank sum of several cluster validation metrics, with the 5-cluster model achieving the best overall performance across metrics (Silhouette Score: 0.42, Calinski-Harabasz Index: 118.76, Davies-Bouldin Score: 0.72, AIC: 856.29) (figure S6).

### Neurobiological Cluster Characterisation

Examining the means and distributions of neuroimaging outcomes across modalities revealed distinct patterns (Fig. 5), which we statistically confirmed using a pairwise comparison test followed by FDR correction. Cluster 5 stands out with consistently high values across all modalities and shows highly significant differences compared to all other groups in mean GMV (all FDR*_p_* < .001). Cluster 3 exhibits the highest FA values, significantly higher than Cluster 2 (FDR*_p_* <.001), but also as Clusters 1 (FDR*_p_*< .001) and 4 (FDR*_p_*= 0.01).

**Figure 5:**
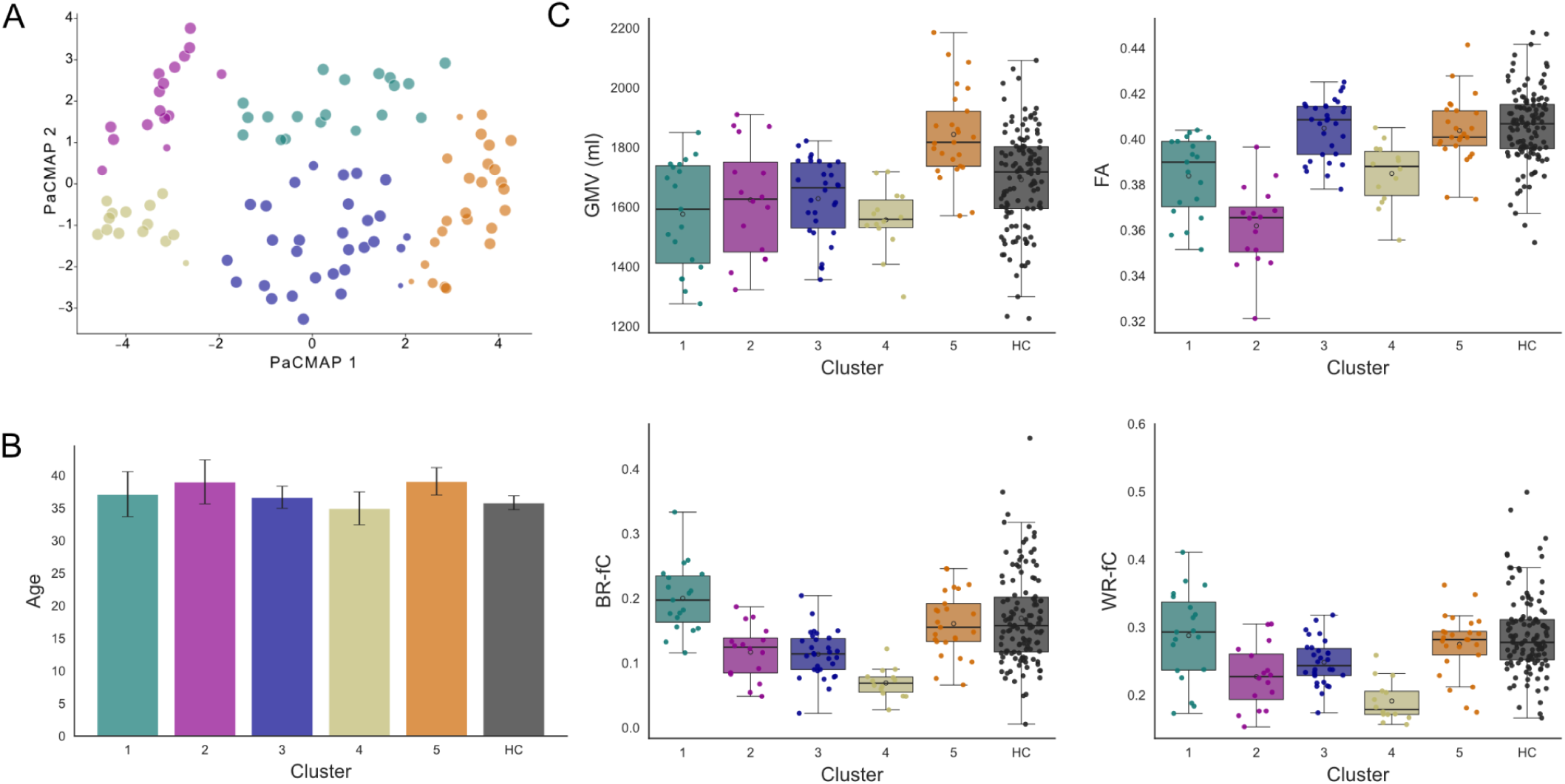
Cluster Structure and neuroimaging outcomes. **A)** Two-dimensional Cluster structure. While the clusters did not differ in confounding variables such as age **(B)**, they differed significantly in mean neuroimaging outcomes **(C)**. PacMAP = Pairwise Controlled Manifold Approximation and Projection.

Cluster 1 has the highest BR-fC and WR-fC, with significantly higher BR-fC compared to Clusters 2 - 4 (FDR*_p_* < .001) and to Cluster 5 (FDR*_p_* = 0.025).

In contrast, Cluster 2 and Cluster 4 display low mean values across modalities, with Cluster 2 particularly low in FA and Cluster 4 notably reduced in functional connectivity. In marked contrast to other clusters and regions, Cluster 2 also displayed sharp GMV reductions in the bilateral rostral Thalamus and medial area 10 (Fig. 6B).

**Figure 6:**
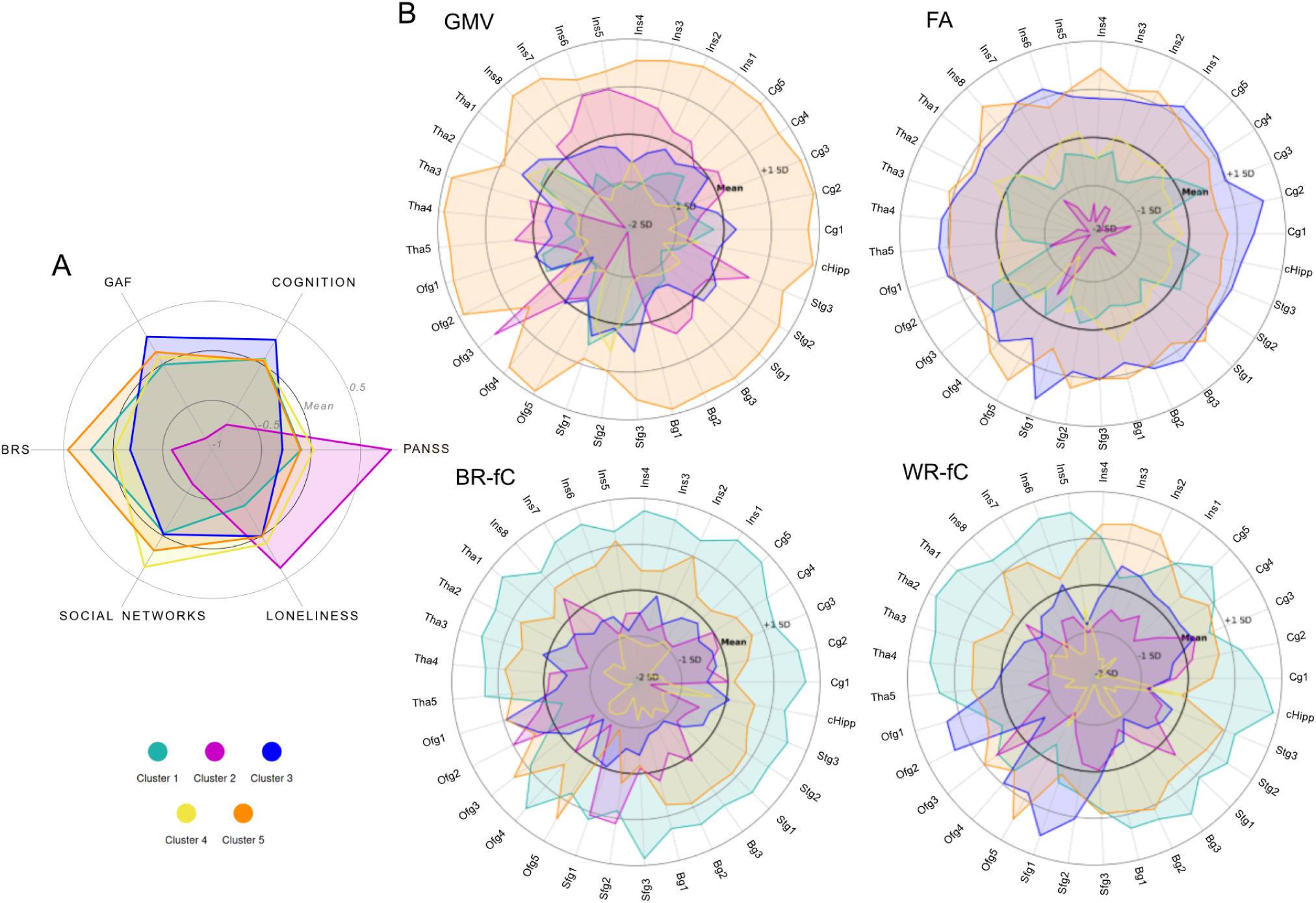
Cluster Characterisation. **A)** Clinical Scores. **B)** The 33 meta-analytic brain regions across modalities (Top left: GMV=Gray Matter Volume; Top right: FA=Fractional Anisotropy; Bottom left: BR-fC=Between Region functional connectivity; Bottom right: WR-fC=Within Region functional connectivity). BRS=Brief Resilience Scale. GAF=Global Assessment of Functioning. PANSS=Positive and Negative Syndrome Scale. The names of the regions as listed in the Brainnetome Atlas are displayed in Fig. 2.

Regarding mean FA, all pairwise comparisons were significant except between Cluster 1 and Cluster 4, and between Cluster 3 and Cluster 5. Regarding BR-fC, all group comparisons were statistically significant except between Clusters 2 and 3, with the strongest differences between Cluster 1 and other clusters. A similar, albeit less pronounced, pattern was observed in WR-fC, where Cluster 1 had significantly higher values than Clusters 2, 3, and 4 (FDR*_p_* < 0.05), but not significantly different from Cluster 5. Fig. 6 shows the distribution of GMV, FA and BR-fC across all single regions.

### Clinical and Sociodemographic Cluster Characterisation

The distribution of patients across clusters was as follows: 19, 16, 30, 14, and 25 in Clusters 1 through 5, respectively. The youngest mean age was observed in Cluster 4 (34.9 years) and the oldest in Cluster 5 (39.1 years), with no significant age differences between clusters (lowest p = 0.815). Crucially, despite age being a well-known factor influencing volumetric brain loss, our oldest cluster (Cluster 5) showed the highest GMV and the second-highest FA values, excluding age as primary driver of the cluster structure. Other potentially confounding factors, including education, BMI and chlorpromazine (CPZ) equivalent dosage, also showed no significant differences between clusters (see table S7).

When comparing clinical scores across clusters, we found that Cluster 2, which exhibited sharp reductions in FA and a unique pattern of GMV distribution, was consistently more burdened across clinical metrics (Fig. 6 A). The clusters differing significantly from Cluster 2 varied depending on the clinical measure, a pattern mirroring the distribution of neuroimaging outcomes. For example Cluster 5 had significantly higher GMV and resilience compared to Cluster 2, while Cluster 3 had significantly higher FA and GAF compared to Cluster 2.

The most substantial clinical difference emerged in GAF scores between Clusters 3 (58.9 ± 12.6) and 2 (44.3 ± 5.8) (d = 1.37, CI [2.03, 0.7], FDR*_p_* < .001). Samara et al. describe a GAF of 45 as serious impairment, a GAF of 65 as moderate difficulty (10). Furthermore, Cluster 2 showed a markedly higher PANSS total score (63.9 ± 16.4) compared to Cluster 3 (47.7 ± 14.7) (d = 1.06, CI [0.42, 1.71], FDR*_p_* = 0.013) and was the only cluster with a score exceeding the threshold proposed for a “mildly ill” classification (PANSS >58) (11). This pair of clusters was also the only one to differ significantly in cognition (d = 0.96, CI [1.6, 0.32], FDR*_p_* = 0.033).

In terms of resilience, measured by the Brief Resilience Scale (BRS), Cluster 5 (BRS = 3.16 ± 0.67) significantly outperformed Cluster 2 (BRS = 2.33 ± 0.71) (d = 1.2, CI [0.47, 1.93], FDR*_p_* = 0.014). Notably, the mean difference in BRS scores between the high- and low-resilience patient clusters was twice as large as the difference between our high-resilience patients cluster and two samples of undergraduate students (mean score students: 3.55 ± 0.72) (12), indicating a significant disparity in this measure between our patient groups. The PANSS total score between Cluster 2 and Cluster 5 differed also significantly (d = 0.89, CI [0.23, 1.55], FDR*_p_* = 0.044).

Further differences emerged in the loneliness and social networks scales, highlighting varying social support patterns across clusters. The loneliness scale showed a moderate effect size (d = 0.65, CI [1.39, 0.05]) when comparing Cluster 1 (mean = 6.47

± 1.95) and Cluster 2 (mean = 8.15 ± 3.16), with a p-value approaching significance (p=0.075). However, this scale lacks a neutral midpoint, meaning that a total score of 6 typically indicates patients answered “rarely” (score of 2) to all three questions, while a score of 8 would suggest that most responses shifted to “often” (score of 3), indicating that even small differences may reflect meaningful shifts in perceived loneliness.

Interestingly, while the Lubben Social Network Scale did not show significant differences in the family subscale between these clusters, the friends subscale revealed a strong effect (d = 1.15, CI [1.92, 0.36], FDR*_p_* = 0.015), suggesting that the trending effect in loneliness might be associated with a lack of social networks beyond family members.

For a detailed overview over clinical scores we refer to table S8.

Furthermore, the distribution of 8 patients with history of electroconvulsive therapy (ECT) across clusters differed significantly (Chi-square p = 0.031). Notably, Cluster 3 as the largest cluster (n = 30) had no patients with ECT history, while Cluster 2 and 4 as the smallest clusters each had three patients. With ECT being used in particularly treatment-resistant cases, this further highlights the clinically relevant information captured by the neuroimaging-derived clusters.

While there were many differences in clinical scores between Cluster 2 and other subgroups, there were no significant clinical differences between the remaining Clusters.

### Genetics, childhood adversity and illness duration: Differences between clusters

The PRS for ICV was significantly higher in Cluster 5 compared to Cluster 4 (d = 1.21, CI [2.01, 0.41], FDR*_p_* = 0.034), Cluster 3 (d = 1.04, CI [1.69, 0.4], FDR*_p_* = 0.034) and healthy controls (d = 0.66, CI [1.16, 0.16], FDR*_p_* = 0.049). This aligned with Cluster 5 having significantly greater GMV than all other clusters, highlighting a potential link between polygenic load for intracranial volume and structural brain differences. The correlation between the PRS for ICV and GMV across all patients was moderately positive (r = 0.48; p <0.001) (Fig.7). No significant differences were observed in regional brain PRS scores or clinical PRS scores across clusters.

**Fig. 7:**
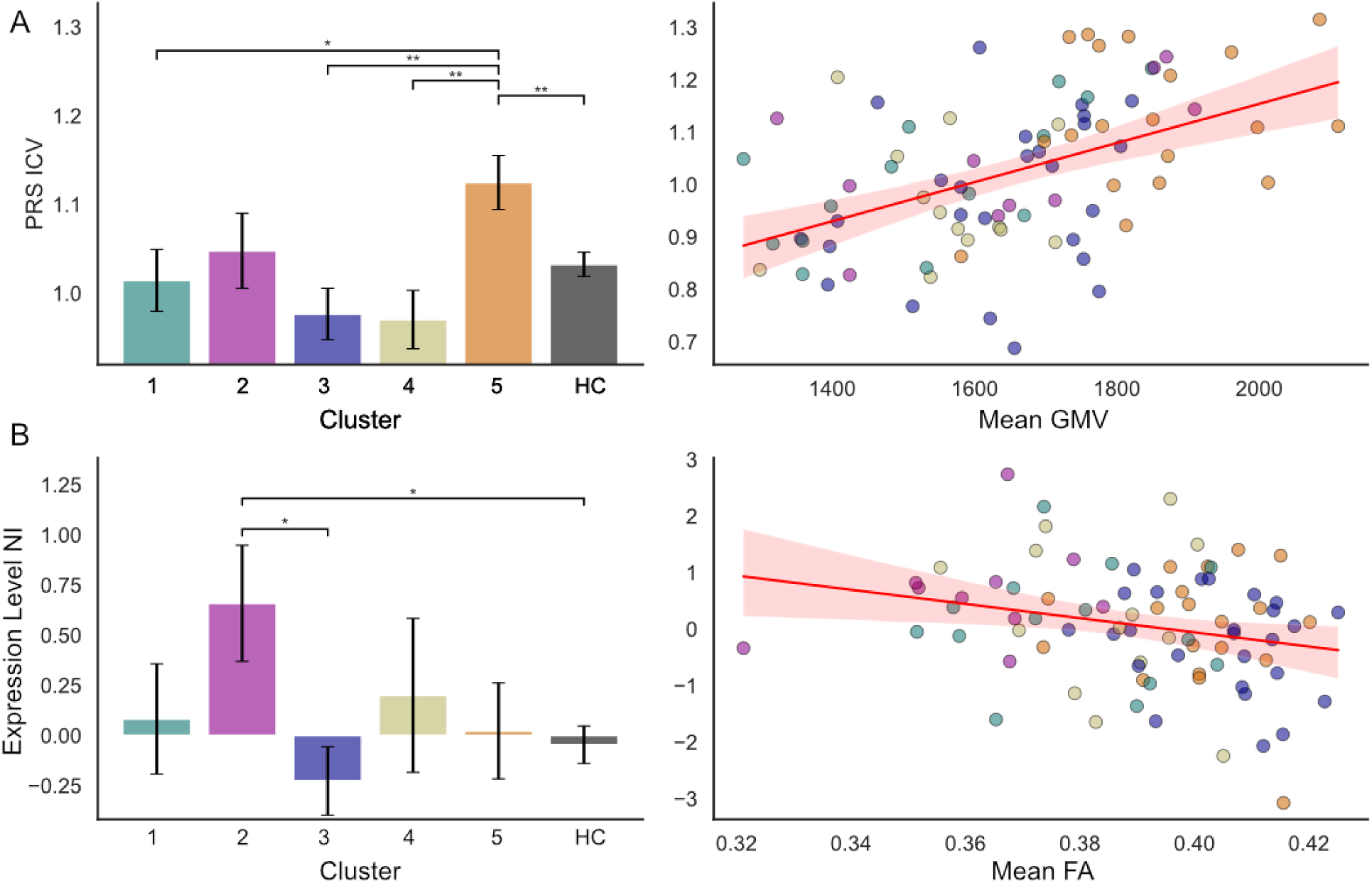
Brain-Genetics associations. **A) Left:** Cluster 5 showed an increased polygenic risk score for Intracranial Volume (PRS ICV), corresponding to its high Gray Matter Volume (GMV). **Right:** The correlation between PRS ICV and mean GMV was significant across all patients (r = 0.478, p = <.001). **B)** Cluster 2 differed significantly in the Neuroinflammation (NI) pathway expression and across all patients the NI pathway correlated with mean Fractional Anisotropy (r = - 0.24, p = 0.027).

For the gene expression pathways, only neuroinflammation showed a significant negative correlation with mean FA (r = - 0.24, p = 0.027). Furthermore, the group comparison revealed significant differences comparing Cluster 2 with Cluster 3 (d = 1.01, CI [1.8, 0.21], p = 0.023) and with healthy controls (d = 0.73, CI [1.4, 0.07], p = 0.042) (Fig. 7).

Additionally, the expression levels for the myelin pathway differed significantly between Cluster 2 and 3, 2 and 1, and Cluster 2 and healthy controls (p = 0.025, p = 0.043 and p = 0.046 respectively), with comparable effect sizes. The correlation between this pathway and mean FA was not significant (r = 0.13, p = 0.251) (figure S5). None of the associations between pathway scores and cluster differences or neuroimaging outcomes survived FDR correction.

Neither the CTS sum scores nor the trauma subdomains showed significant differences between patient clusters. Likewise, there was no significant difference in illness duration, with mean values ranging from 7.8 to 12.1 years (see table S8).

### Brain - Symptom - Interactions

To further investigate the relationship between symptoms and our ROIs, Pearson correlations of each clinical variable with each of the 33 brain regions in 4 modalities were calculated. FDR correction was applied for each variable separately across 133 correlations. All correlations exceeding the threshold of a weak relationship (r ≥ 0.2, p < 0.05) are displayed in the supplementary figure 7A. Interestingly, the correlations between neuroimaging modality and clinical score varied widely: GAF was predominantly positively correlated with FA in several regions, size of social networks was predominantly positively correlated with GMV, and perceived loneliness was negatively correlated with BR-fC.

Among the strongest correlations, a predominance of multimodal insular values emerged: Higher GAF was associated with higher FA in the bilateral ventral granular and dysgranular insula (r = 0.31, p <.001). Higher loneliness was associated with lower BR-fC in the left ventral granular and dysgranular insula (r = −0.36, p <0.001), left dorsal dysgranular insula (r = −0.34, p <.001), right dorsal agranular insula (r = −0.33, p <.001) and lateral area 12/47 (orbital gyrus) (r= −0.33, p<.001).

Regions with the strongest correlations with clinical scores often contributed most to neuroimaging-based variance. For example, the ventral granular and dysgranular insula consistently exceeded the r ≥ 0.2 threshold across modalities and clinical scores, while also correlating strongly with both PaCMAP projections, driving cluster differentiation.

Notably, this is not a predetermined relationship, as a variable can drive cluster differentiation due to high variability without being clinically meaningful.

Next, we asked if the observed brain-symptom associations hold true across bootstrap runs. We created multimodal, questionnaire-specific composite neuroscores by aggregating all neuroimaging values exceeding the threshold of a weak relationship for a given clinical variable (r = > 0.2, p = < 0.05), standardizing, and averaging them. The GAF-composite (r = 0.33; p = 0.002), Social Network-composite (r = 0.33; p = <.001), Resilience-composite (r = 0.3; p = 0.005) and Loneliness-composite (r = - 0.3; p = 0.005) neuroscores exhibited the highest correlations with their respective clinical variables. Correlations between the magnitude of clinical differences and the respective composite scores emerged across bootstrap runs, the most notable being a correlation of 0.47 (p <.001) between the GAF and its neuroscore, consisting mostly of FA values (see figure S7B). This means that the greater the difference in FA was between clusters, the greater their respective difference in GAF. To assess the specificity of the GAF neuroscore, we examined its correlations with unrelated clinical variables. These analyses revealed much weaker relationships, with loneliness (r = 0.11, p <.001) and resilience (r = 0.14, p <.001) showing only nominal significance. While some correlation across clinical scores is expected—clusters with higher clinical scores are likely to exhibit overall higher volumetric outcomes—the comparatively strong correlation of the GAF neuroscore with the GAF clinical variable suggests a more specific relationship between them.

### Cluster stability and pattern robustness

First we compared our cluster validation metrics against null distributions generated from synthetic datasets. For details regarding this technique we refer to the methods section. Unlike the values obtained in previous studies (13), all indices here were significant under their respective null distributions (p <0.001), with no single simulation achieving the CH or silhouette score we see for our real clusters (see Figure S6).

Then, based on our real data, we performed 10 000 bootstrap iterations. The mean silhouette score for a 5-cluster structure was 0.39, with a range from 0.22 to 0.49. However, when letting the algorithm choose flexibly between 2 and 6 clusters, optimising for the best silhouette score as we have done in our complete cohort, the lower end improves substantially, with a range of 0.35 to 0.57 and a mean of 0.44. These results point to the overall ability of our workflow to produce well-separated multimodal clusters.

Finally, given that PaCMAP has not yet been used in neuroimaging, we rerun our analysis, with Uniform Manifold Approximation and Projection for Dimension Reduction (UMAP) as a widely adopted nonlinear DR technique serving as a further benchmark against which to evaluate our findings. Clustering comparison metrics revealed moderate to strong agreement between the two approaches, with Cohen’s Kappa values for specific cluster pairs ranging from 0.64 to 0.75. While both DR techniques identified consistent neuroimaging-clinical relationships and preserved the core structure of the clusters, PaCMAP based clustering was able to differentiate the most burdened patients by correctly identifying underlying differences in neuroimaging patterns. We provide a detailed comparison of PaCMAP-based clustering and UMAP-based clustering in the supplementary material (figures S9-S11).

## Discussion

The heterogeneity of SSD is a major challenge to the discovery of core biological mechanisms. To address this, we developed a workflow for multimodal neuroimaging clustering, uncovering robust and clinically meaningful subgroups within an otherwise stable cohort of SSD patients.

First, we confirmed reduction in GMV in meta-analytically derived ROIs, replicating meta-analytic findings in 30/33 regions in the internal and in 22/33 regions in the external comparison. The lower external replication rate likely reflects differences in control groups, with external controls stemming from a diverse general population and internal controls reflecting a convenience sample with unusually high educational achievement and unusually low symptom burden.

The identification of 22 significant regions across cohorts underscores the robustness of the meta-analytically selected ROIs. Furthermore, multimodal analysis revealed significant differences in 16 regions across GMV, FA, and functional connectivity metrics, further confirming the central role of these regions in SSD.

Largest effects sizes were observed in the thalamus, anterior cingulate, and insula - key components of the salience network (81,82). Disturbances in this network are thought to contribute to faulty perceptions of salient external and internal events in SSD (83,84).

These findings align with large-scale meta-analyses, with Sha et al. (77) reporting reduced GMV and altered connectivity in the bilateral insula and ACC, while in Goodkind et al (14) the thalamus showed the strongest effects for GMV reductions. Meta-analytic white matter findings have shown lower whole-brain Fractional Anisotropy (FA) and higher mean diffusivity in patients with schizophrenia (17,18). Many of the frequently implicated tracts, such as the corona radiata and the anterior limb of the internal capsule, connect the thalamus to other brain regions, aligning with the prominent thalamic effect sizes observed in FA within our cohort.

Widespread reduced connectivity within and between brain networks is thought to contribute to the characteristic difficulties of SSD patients to maintain an integrated sense of Self (85) and evidence from large studies (86) and meta-analysis (87) points towards reduced thalamo-frontal connectivity and increased thalamus-primary somatosensory cortex connectivity. Given that our ROIs include frontal regions but no regions belonging to the somatosensory cortex, the overall hypoconnectivity we find in patients aligns with these observations.

The predominance of thalamic effects in our cohort further aligns with the thalamic PRS being the only brain PRS to show trending significance in case-control comparisons and showing of all PRS scores the strongest correlations with neuroimaging values, specifically for GMV and FA in controls. Interestingly, recent large-scale studies highlight potential links between thalamic structure and genetic risk factors in SSD. Stauffer et al. (88), using bidirectional mendelian randomization, found that reduced genetically predicted neurite density index (NDI) in the thalamus was associated with increased genetically predicted risk for schizophrenia, suggesting that genetic variants influencing thalamic microstructure may causally contribute to schizophrenia risk. In a study of 81 adolescents with 22q11.2 deletion syndrome, the most common microdeletion in humans and one of the highest known risk factors for psychosis (20-40% of adults carriers of this deletion are diagnosed with a psychotic disorder (89)), thalamic connectivity was clearly altered, pointing further towards a possible relationship between genetics, thalamus and psychotic experiences (90). These findings support the notion that individual genomic background influencing thalamic structure and function may play a role in SSD, potentially contributing to the pronounced thalamic effect sizes in our study.

To address the neurobiological and clinical heterogeneity of SSD, we used PaCMAP-based GMM clustering and identified five clusters with distinct biological profiles. Cluster 1 demonstrated high functional connectivity and low loneliness, Cluster 3 high FA, GAF and cognition scores, Cluster 5 high GMV, resilience and PRS score for ICV. While there were many significant differences between clusters in terms of neuroimaging and genetics, only Cluster 2 stood out with regard to clinical burden. This was associated with significantly reduced FA across regions, low GMV in bilateral rostral thalamus and medial area 10, increased expression of neuroinflammation and decreased expression of myelin associated genes. We confirmed associations between neuroimaging outcomes and clinical scores by correlation analysis across all patients and bootstrapping highlighted the correlation between FA and GAF, a pattern aligned with previous literature. Yamada et al. reported positive associations between FA and GAF in two independent cohorts (91), and Kelly et al. (17) found trending negative correlations between symptom severity and FA in specific white matter tracts. Additionally, GAF scores have been shown to correlate positively with a range of cognitive measures (92,93), a relationship mirrored in our clusters. Noteworthy, a large multisite harmonization study demonstrated that widespread white matter disruptions in schizophrenia significantly affect cognitive functioning (94), while a recent longitudinal study of major depressive disorder involving 418 patients and 462 controls found that individuals with cognitive decline exhibited concurrent reductions in white matter integrity (95). Furthermore, higher baseline FA values in SZ have been shown to predict cognitive training-induced improvements in attention and vigilance (96).

Both the axon itself and the myelin sheath contribute to FA (97). Through myelin formation, oligodendroglial cells contribute to the propagation velocity of electrical impulses along nerve fibers (98). A dysfunction on this level would influence connection between brain regions, impacting higher-order functions such as cognition and behavior (95). Interestingly, both numerical reduction and altered metabolism of oligodendrocytes were found in schizophrenia (99). Downregulation of myelin structural proteins and reduction of mRNA levels of various oligodendroglial lineage transcription factors were found in postmortem analysis (100,101). The effect of oligodendrocyte gene variants on cognition was found to be mediated via the integrity of white matter tracts (102) and a recent hypothesis suggests that disturbed maturation of oligodendrocyte precursor cells in a subgroup of patients leads to cognitive deficits (103). Furthermore, a meta-analysis showed neuroinflammatory mechanisms to be associated with white matter pathology in schizophrenia (104) and a gene expression-based clustering approach identified a subgroup enriched for immune-related pathways and genetic profiles previously associated with reduced fractional anisotropy (67). These findings align with our Cluster 2 showing remarkably reduced white matter integrity, increased expression of neuroinflammation related genes and decreased expression of myelin related genes. However, given our sample size and that genetic associations do not survive FDR correction, the findings should be interpreted with caution and further studies with larger cohorts are needed to confirm these preliminary observations.

Small social networks and high loneliness are common in SSD and patients often cite “improved relationships” as a major treatment goal (105). Limited data exists on the relationship between loneliness and functional connectivity, but negative correlations have been reported in healthy controls (106,107). This aligns with Cluster 1 having distinctively high functional connectivity and low perceived loneliness.

Positive associations between GMV and resilience, as exhibited by our Cluster 5, remain underexplored too. Notably, a six-year follow-up study of adolescents at ultra-high risk for psychosis (UHR) identified resilient and non-resilient groups, with the resilient group showing significantly greater cortical volume and thickness at baseline (108). Another study of adolescents with and without persistent attenuated psychotic symptoms (APS) found that those with APS at follow-up exhibited reduced baseline GMV, despite being indistinguishable in clinical and demographic variables at study onset (109). These studies provide an intriguing context for interpreting Cluster 5.

Despite being the oldest cluster, it shows higher GMV than all other clusters, likely reflecting the influence of its significantly higher PRS score for ICV. This would suggest that Cluster 5 may have been distinguishable by high GMV decades earlier, with its elevated resilience and favorable clinical profile fitting the trajectory described in previous studies. These findings suggest that PRS for ICV could indirectly be tapping into resilience mechanisms.

It is important to note that the patterns we found are not necessarily specific to SSD. For instance, Kochunov et al. demonstrated that cognitive performance correlates with FA in both patients and controls, with mediation effects along the FA to processing speed to working memory pathway observed in both groups (110). However, crucially, white matter regions most affected in schizophrenia exhibited the strongest mediation effects, underscoring their particular relevance to SSD. Similarly, the correlation between resilience and gray matter volume has been studied in non-psychiatric cohorts (111). It is possible for there to be complex relationships between brain patterns and clinical variables that are amplified in but not specific to mental health disorders. It is likely that this amplification is more pronounced in subgroups of patients due to equally complex gene-environment interactions.

Finally, the starkly reduced white matter integrity of Cluster 2 reminds of the different but complementary framework of the neurodegenerative model, in which neurodegenerative and neurodevelopmental processes are thought to affect SSD patients to varying degrees, reflecting heterogeneous genetic, epigenetic and environmental influences (112).

Contrary to the assumption of a simple dichotomy separating highly burdened from less burdened patients, our data-driven approach reveals a nuanced structure with five distinct subgroups. To identify these subgroups, our workflow for multimodal clustering leverages meta-analyses-derived ROIs as the most critical hypotheses and ignores other regions, thereby enhancing statistical power and minimizing the risk of false positives (113,114). To integrate multiple modalities (MRI, DTI, and fMRI), we utilized PaCMAP, which, to our knowledge, has not yet been applied in neuroimaging. PaCMAP outperforms other dimensionality reduction techniques in preserving both local neighborhoods and global structures, as demonstrated in benchmark datasets and recent transcriptomics applications (39)(40).

Replication remains a primary concern, and this study was designed with that in mind. By focusing on regions consistently implicated in SSD across meta-analyses encompassing tens of thousands of patients, we aim to provide a foundation that can be generalised to other cohorts. While the exact number and structure of clusters will inevitably vary across samples, multimodal subgroups with reproducible brain-symptom-genetics patterns could significantly refine our understanding of SSD nosology, enhance predictive capabilities, and lay the groundwork for more tailored treatment strategies. However, the novel analytic approach introduced here must be tested in larger, independent datasets before further conclusions can be drawn.

Comprehensive characterization of psychiatric populations is critical to uncovering robust brain-symptom relationships. We consider our reliance on limited clinical measures one of the main limitations of this study. For example, the Childhood Trauma Screener (CTS) attempts to capture sensitive information regarding participants’ childhood in just five questions and lacks validation for psychotic disorders. Witt et al. (115), in a large population survey, conclude the CTS may be useful as a control variable, but is “not sufficient for a comprehensive assessment of child maltreatment.” Given the strong link between childhood adversity and psychiatric outcomes (116), with a recent meta-umbrella systematic review finding that childhood adversities “accounted for about two-fifths (38%) of global cases of schizophrenia spectrum disorders” (117), more thorough tools should be used in future studies. Similarly, while we included the three-item loneliness scale, the 20-item version has shown stronger psychometric properties in SSD populations (118).

The cross-sectional nature of this study inherently limits the scope of interpretation. Without longitudinal data we do not know the temporal stability of the observed patterns. For example, it is well conceivable that with changes in social networks and perceived loneliness functional connectivity would change and hence cluster assignment.

Similarly, if we think of at least some SSD cases as a progressing condition, we would expect cluster profiles to change even in the hypothetical case of all other factors remaining unchanged. If true, the dynamic nature of cluster assignments should be studied in terms of the right moment for therapeutic interventions.

## Conclusion

We could replicate gray matter volume reductions in most of meta-analyses derived regions and found consistent alterations across neuroimaging modalities, confirming the central importance of these regions in SSD. Despite examining an overall stable cohort of patients, our multimodal clustering workflow, grounded in meta-analyses, successfully identified a distinctly burdened subgroup. The meaningful convergence across clinical, neuroimaging, and genetic domains, reinforces the validity of these clusters and highlights the potential of multimodal methodologies to disentangle the heterogeneity associated with mental illness.

However, given the novelty of this workflow and our modest sample size, external replications are needed. If future research uncovers comparable patterns, multimodal stratification could significantly refine subgroup characterization in mental illness, ultimately informing more tailored treatment strategies and advancing precision psychiatry.

## Supplementary material

### Part A: Case-Control

**Table S1:**
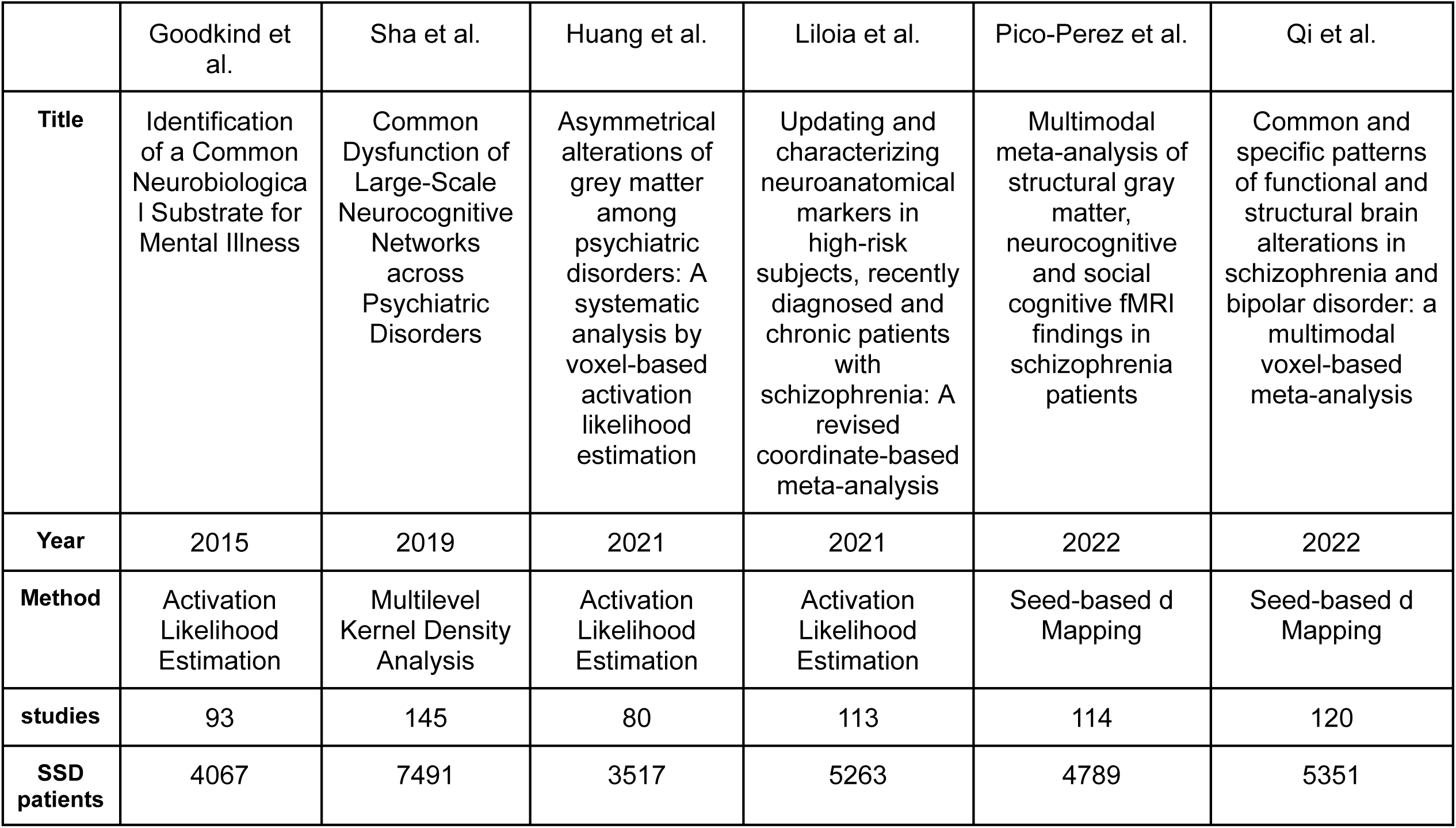
Included Meta-analyses. Goodkind et al., *Jama Psychiatry* 2015. Sha et al., *Biological Psychiatry* 2019. Huang et al., *Progress in Neuropsychopharmacology & Biological Psychiatry* 2021. Liloia et al., *Neuroscience and Biobehavioral Reviews* 2021. Pico-Perez et al., *Psychological Medicine* 2022. Qi et al., *Journal of Psychiatry & Neuroscience* 2022. For ROIs visualisation see Fig. S1.

|  | Goodkind et al. | Sha et al. | Huang et al. | Liloia et al. | Pico-Perez et al. | Qi et al. |
| --- | --- | --- | --- | --- | --- | --- |
| **Title** | Identification of a Common Neurobiological Substrate for Mental Illness | Common Dysfunction of Large-Scale Neurocognitive Networks across Psychiatric Disorders | Asymmetrical alterations of grey matter among psychiatric disorders: A systematic analysis by voxel-based activation likelihood estimation | Updating and characterizing neuroanatomical markers in high-risk subjects, recently diagnosed and chronic patients with schizophrenia: A revised coordinate-based meta-analysis | Multimodal meta-analysis of structural gray matter, neurocognitive and social cognitive fMRI findings in schizophrenia patients | Common and specific patterns of functional and structural brain alterations in schizophrenia and bipolar disorder: a multimodal voxel-based meta-analysis |
| **Year** | 2015 | 2019 | 2021 | 2021 | 2022 | 2022 |
| **Method** | Activation Likelihood Estimation | Multilevel Kernel Density Analysis | Activation Likelihood Estimation | Activation Likelihood Estimation | Seed-based d Mapping | Seed-based d Mapping |
| **studies** | 93 | 145 | 80 | 113 | 114 | 120 |
| **SSD patients** | 4067 | 7491 | 3517 | 5263 | 4789 | 5351 |

#### Inclusion criteria

Published between 2015 and 2024, using a case-control design with a minimum of 1,500 patients with schizophrenia-spectrum disorders, adults only, with whole-brain results for GMV reported in MNI space.

The **search** was performed in pubmed and scopus using the following search terms: Schizophrenia AND meta-analysis AND gray matter; schizophrenia AND systematic review AND gray matter; schizophrenia spectrum disorders AND meta-analysis AND gray matter; schizophrenia AND meta-analysis AND voxel-based morphometry; schizophrenia spectrum disorders AND meta-analysis AND voxel-based morphometry.

### Magnetic Resonance Imaging (MRI)

#### ROIs of meta-analyses

**Fig. S1:**
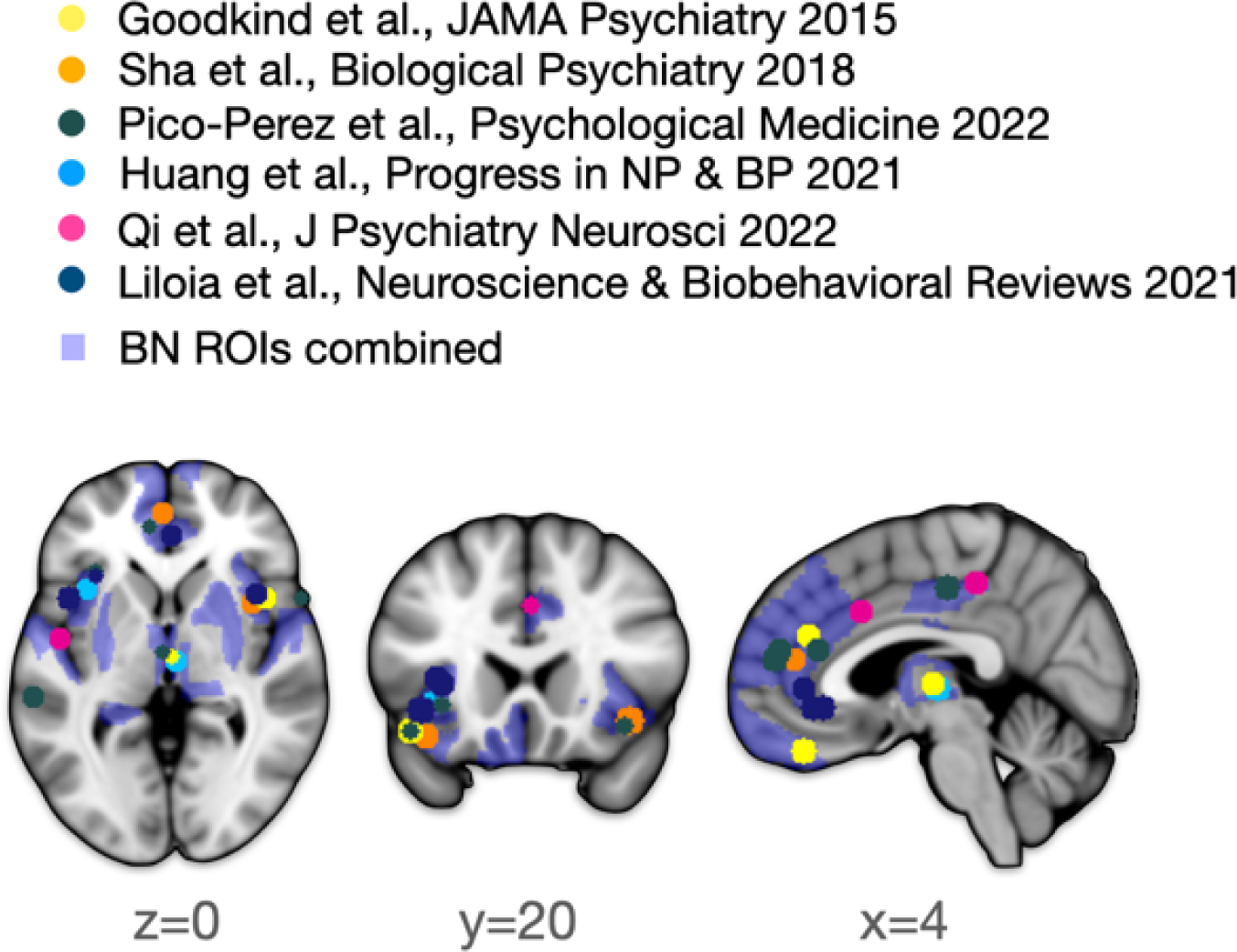
The meta-analytic ROIs of Goodkind et al. are shown in yellow, of Sha et al. in orange, of Pico-Perez et al. in dark green, of Huang et al. in light blue, of ZhangZhang et al. in pink and of Lilia et al. in dark blue, underlaid by the Brainetome Atlas (REF) regions, where at least two meta-analytic ROIs were localized. *The MNI standard template was used for visualization*.

#### MRI Data Acquisition

The scanning protocol of the CDP study was based on the HCP Ageing sequences (Harms et al., 2018). The T1-weighted MPRAGE sequence was acquired with an isotropic voxel size of 0.8 mm. Imaging parameters include a repetition time (TR) of 2500 ms, an echo time (TE) of 2.22 ms and an inversion time (TI) of 1000 ms, flip angle of 8 degrees. A total of 208 sagittal slices were acquired, and GRAPPA acceleration was applied with a factor of 2. The T2-weighted SPACE protocol was used to capture isotropic voxels at a resolution of 0.8 mm. It utilized a TR of 3200 ms and a TE of 563 ms, with a variable flip angle mode to optimize tissue contrast. This sequence captured 208 sagittal slices and included GRAPPA acceleration with a factor of 2. B0 and B1 shimming were applied to improve image uniformity. Resting-state fMRI (rsfMRI) was acquired with an isotropic voxel size of 2.0 mm. The protocol was characterized by a TR of 800 ms and a TE of 37 ms, with a flip angle of 52 degrees. The imaging volume consisted of 72 transversal slices, captured with a multi-band acceleration factor of 8 to enable high temporal resolution. Fat suppression was applied using a fat saturation method to enhance contrast in functional imaging. The diffusion MRI protocol was employed to acquire high-resolution isotropic voxels at 1.6 mm, with a TR of 5500 ms and a TE of 99.2 ms. This protocol captured 90 transversal slices and used a multi-band acceleration factor of 3 to enhance acquisition speed. A monopolar diffusion scheme with 95 directions and a maximum b-value of 2000 s/mm² was applied, with fat suppression included and GRAPPA acceleration disabled to maintain image quality.

#### Multimodal MRI Data Processing

##### Grey Matter Volumes

T1-weighted images, functional EPI images, and DWI images were quality-controlled using the automated quality control software MRIQC (51) combined with visual inspection of all raw images.

The T1-weighted images were processed using the Neuromodulation and Multimodal NeuroImaging software (NAMNIs) version 0.3 (52). NAMNIs is mostly based on FSL and includes a reorientation to the standard space, brain extraction, segmentation, masking, a combination of linear and non-linear registration, and a warping of a brain atlas of choice in the native space of the subjects. As the final output, we extracted gray matter volumes of the meta-analytically derived regions defined by the Brainnetome atlas.

##### Resting State fMRI (rs-fMRI)

Functional EPI images were preprocessed using fMRIPrep version 23.0.1 (53) (see the fMRIPrep documentation for the processing steps included: https://fmriprep.org/en/stable/). Preprocessed images were smoothed (FWHM = 6mm) and denoised using the global signal, cerebrospinal fluid signal, white matter signal, and the extracted noise components from Automatic Removal Of Motion Artifacts based on independent component analysis (ICA-AROMA) as regressors in the *clean_img* from the *image* module in Nilearn (https://nilearn.github.io/stable/modules/image.html).

During denoising, the data was also band-pass filtered (0.01 - 0.1 Hz), detrended, and standardized. The subject-specific denoised BOLD mean time series of the meta-analytically derived regions and of all voxels within these regions were extracted using the *maskers* module from Nilearn (https://nilearn.github.io/stable/modules/maskers.html). Two types of functional connectivity were computed from these time series using the *connectome* module from Nilearn (https://nilearn.github.io/stable/modules/connectome.html): First, between the mean time series of the regions, resulting in a 31 x 31 functional connectivity matrix.

This matrix was averaged column-wise, while ignoring the self-connection of each region, to receive a single functional connectivity score indicating the average functional connectivity between each region and all other regions of interest. Second, functional connectivity between all voxels within each region of interest was computed and the upper triangle of the resulting connectivity matrix was averaged to one score reflecting the functional connectivity strength within each region.

##### Diffusion Tensor Imaging

DTI data were processed using tract-based spatial statistics (TBSS) tools in version 6.0.7.8 of the FMRIB Software Library (FSL). The Brain Extraction Tool was used to generate a binary mask from non-diffusion-weighted data. Diffusion tensor and associated parameters—including fractional anisotropy (FA), mean diffusivity, axial diffusivity, and radial diffusivity—were calculated using the DTIFIT tool in FSL. Nonlinear transformation and affine registration were applied to normalize all FA data into standard space using the FNIRT tool. Normalized FA images were averaged to create a mean FA image, and a mean FA skeleton was generated by identifying the central tracts common to all subjects. The threshold for the mean FA skeleton white matter mask was set between 0.2 and 0.8 to exclude voxels containing gray matter or CSF. The voxel size was set to 1 x 1 x 1 mm in Montreal Neurological Institute (MNI) space.

Each subject’s FA map voxel values were projected onto the skeleton by identifying the local maxima perpendicular to the skeleton. This step ensures each subject’s FA data aligns with the relevant part of the skeleton, correcting for residual misalignment after initial nonlinear registration. Finally, we used only those tracts that passed through our meta-analytical BN ROIs and used these FA values for the subsequent statistical analyses.

### Electroencephalography

#### EEG recording

Resting-state EEG was recorded with 32 electrodes using a BrainAmp amplifier (Brain Products, Germany) at 1000 Hz sampling rate, referenced to Cz, and with impedance below 5 kΩ. Electrodes followed the International 10/20 system. The 10-minute recording included 5 minutes with eyes closed and 5 minutes with eyes open, separated by an event marker. For 11 participants lacking markers, the first and last 3.5 minutes were assigned as eyes-closed and eyes-open sessions. Participants were instructed to remain calm and relaxed.

#### EEG Preprocessing

EEG data were preprocessed using a modified pipeline from Adams et al. (54) in MATLAB (The Mathworks Inc.) with EEGLAB v2022.0 (55) (https://sccn.ucsd.edu/eeglab/), separately for eyes-open and eyes-closed conditions. Data were re-referenced to TP9/TP10, downsampled to 256 Hz, filtered (1–70 Hz), and notch filtered (49.5–50.5 Hz), then segmented into 6-second epochs. Artifacts were removed through a multi-step process: epochs exceeding ±5 SD were excluded, as were epochs with linear trends (max slope=5, min R^2^=0.7), low (<2 Hz; within the power threshold of -50 to 50 dB) or high (20–40 Hz; within the power threshold of -100 to 25 dB) frequencies, or bad channels based on power spectrum (-4 to 6 std), kurtosis (-7 to 15 std), and joint probability (-9 to 7 std) thresholds. If >50% of epochs were rejected, channel rejection was prioritized, followed by epoch rejection with the initial data.

Datasets were discarded if >50% of data or >20% of electrodes were removed. Next, Independent Component Analysis (ICA) was applied with artifact components removed via Multiple Artifact Rejection Algorithm (MARA). A second round of rejection with adjusted thresholds (power spectrum [-6 to 5 std], kurtosis [-6 to 9 std], and joint probability thresholds [-7 to 7 std]) was applied, followed by channel interpolation, and data were converted back to continuous format for analysis.

#### Source Localization

Source localization was conducted in Brainstorm (Version: April-2024) (56) (https://neuroimage.usc.edu/brainstorm) after EEG preprocessing. Anatomical data processed with FreeSurfer v7.3.2 (57,58) (http://surfer.nmr.mgh.harvard.edu/) were imported into Brainstorm, with the cortical surface set to 15,000 vertices for each subject. EEG electrode positions were aligned using the default ICBM 152 10-20 template and projected to the scalp. A subject-specific three-layer BEM head model (1922 vertices per layer for scalp, skull, and brain) was created with default conductivity values (1, 0.0125, 1) and 4 mm skull thickness. The head model was computed with OpenMEEG, using the cortex surface as the source space. Noise covariance was based on an identity matrix, while data covariance was calculated over the entire recording window to capture signal variability. Source estimation used the Linearly Constrained Minimum Variance (LCMV) beamformer (59) with unconstrained dipole orientations, extracting time-series data from 17 ROIs, excluding dlPu ROIs, which failed to be reliably imported as scouts for over 40% of subjects.

#### Power Spectrum Density

Power Spectrum Density (PSD) was calculated for time-series of each ROI using the Welch method with 6-second windows and 3-second overlap in MNE-Python (v1.8.0) (60). Absolute power was extracted across six frequency bands: delta (1-4 Hz), theta (4-8 Hz), alpha (8-12 Hz), beta (12-30 Hz), gamma1 (30-50 Hz), gamma2 (50-70 Hz), low-frequency (1–12 Hz), and high-frequency (12–70 Hz).

**Table S2:** Gray Matter ROIs replication internal cohort.

| Region | HC Mean | P Mean | HC-P Diff | FDR p | Cohen's d | Cohen's d CI |
| --- | --- | --- | --- | --- | --- | --- |
| rTthaR | 818.9 | 719.3 | 99.6 | <0.001 | 0.46 | [0.22, 0.7] |
| A32pR | 1497.2 | 1412.0 | 85.2 | <0.001 | 0.45 | [0.21, 0.69] |
| rTthaL | 894.7 | 814.9 | 79.8 | <0.001 | 0.45 | [0.21, 0.69] |
| TE10TE12R | 2133.1 | 1996.5 | 136.7 | <0.001 | 0.38 | [0.14, 0.62] |
| A11mR | 2130.7 | 2022.2 | 108.5 | <0.001 | 0.37 | [0.13, 0.61] |
| A9mR | 2951.8 | 2822.5 | 129.4 | <0.001 | 0.37 | [0.13, 0.61] |
| A10mR | 2517.6 | 2390.6 | 127.0 | 0.007 | 0.34 | [0.1, 0.58] |
| A1247oL | 1625.8 | 1548.5 | 77.3 | <0.001 | 0.34 | [0.1, 0.58] |
| A32sgR | 1708.3 | 1642.0 | 66.3 | <0.001 | 0.31 | [0.07, 0.55] |
| dlaR | 1052.8 | 1015.3 | 37.5 | <0.001 | 0.3 | [0.06, 0.54] |
| A10mL | 2722.1 | 2603.0 | 119.1 | 0.028 | 0.28 | [0.04, 0.52] |
| dlaL | 1181.6 | 1138.1 | 43.5 | <0.001 | 0.27 | [0.03, 0.51] |
| vlaR | 927.2 | 890.7 | 36.5 | <0.001 | 0.27 | [0.03, 0.51] |
| dldL | 1785.0 | 1718.4 | 66.6 | <0.001 | 0.25 | [0.01, 0.49] |
| dldR | 1390.1 | 1340.4 | 49.8 | <0.001 | 0.25 | [0.01, 0.49] |
| A14mL | 1744.5 | 1688.3 | 56.2 | 0.003 | 0.25 | [0.01, 0.49] |
| A1247IR | 1516.2 | 1468.2 | 48.0 | 0.003 | 0.24 | [0.0, 0.48] |
| cHippL | 2590.9 | 2504.6 | 86.3 | 0.001 | 0.24 | [0.0, 0.48] |
| mPFthaL | 1016.1 | 973.3 | 42.8 | <0.001 | 0.24 | [0.0, 0.48] |
| vidvlgR | 1376.4 | 1335.3 | 41.1 | <0.001 | 0.23 | [-0.01, 0.47] |
| A32sgL | 2481.2 | 2406.7 | 74.6 | 0.001 | 0.22 | [-0.02, 0.46] |
| vlaL | 1071.6 | 1041.9 | 29.7 | <0.001 | 0.22 | [-0.02, 0.46] |
| TE10TE12L | 2308.3 | 2227.3 | 81.0 | 0.003 | 0.21 | [-0.03, 0.45] |
| A4142R | 828.5 | 874.7 | -46.2 | 0.785 | -0.18 | [-0.42, 0.06] |
| A23cL | 2235.4 | 2186.7 | 48.8 | 0.001 | 0.17 | [-0.07, 0.41] |
| A23cR | 2087.0 | 2046.5 | 40.5 | 0.004 | 0.15 | [-0.09, 0.39] |
| vldvlgL | 1204.6 | 1180.4 | 24.1 | 0.001 | 0.14 | [-0.1, 0.38] |
| A13L | 2808.7 | 2889.6 | -80.9 | 0.932 | -0.12 | [-0.36, 0.12] |
| PPthaR | 1064.8 | 1041.8 | 23.0 | 0.005 | 0.12 | [-0.12, 0.36] |
| dIPuR | 1664.2 | 1600.3 | 63.9 | 0.009 | 0.12 | [-0.12, 0.36] |
| dIPuL | 1730.5 | 1679.9 | 50.7 | 0.015 | 0.1 | [-0.14, 0.34] |
| vmPuR | 1070.2 | 1080.7 | -10.6 | 0.124 | -0.04 | [-0.28, 0.2] |
| IPFthaR | 1495.3 | 1488.3 | 7.0 | 0.023 | 0.03 | [-0.21, 0.27] |

### Site-harmonization with the external cohort

Given the moderate sample size and sex ratio imbalance in our original control group, we pooled controls from the young and old cohorts of the Human Connectome Project (HCP) with controls from our own cohort to increase sample size and to externally validate findings. Including some CDP controls helped bridge the gap in age range between otherwise disjoint age ranges of the two HCP cohorts, providing a smoother age distribution. Using a matching algorithm that prioritized HCP controls over CDP controls, we created a balanced, mostly external cohort with identical age and sex distributions between patients and controls. This algorithm selected controls based on exact age (+/- 1 year) and sex matching, allowing for a more representative and demographically balanced control group.

To harmonize imaging data across multiple sites within this expanded, multisite cohort, we applied the ComBat-GAM method, an advanced extension of the ComBat harmonization framework (119). By leveraging generalized additive models, ComBat-GAM enables nonlinear adjustments, allowing us to include both linear and quadratic variables for age to account for nonlinear age effects on brain structure.

The harmonization model was trained exclusively on control subjects, ensuring that the adjustments were based solely on data unaffected by disease pathology. This strategy preserves group differences by reducing site-related variance without inadvertently diminishing patient-specific effects. To capture individual anatomical variability, we also included Estimated Total Intracranial Volume (ICV) as a covariate, controlling for differences in brain size across subjects, in line with the ComBat-GAM methodology (119).

To ensure that our harmonization covered the full age range of our sample, we extended the control data by including synthetic data points at the age range boundaries, allowing the model to generalize effectively to both younger and older participants. These “dummy” points were used solely to stabilize the harmonization model across the entire age distribution.

After training on control data, we applied the harmonization model to align all subjects within a common statistical space, minimizing site-related effects.

To assess the impact of site harmonization, we calculated R² values pre- and post-harmonization for site, sex, and age effects. Harmonization significantly reduced the site effect, with the mean R² across all regions halving to 4.4%. In contrast, R² for sex remained stable at approximately 10%, suggesting that the harmonization effectively targeted site variability without altering sex-related variance. Notably, R² for age effects showed opposite shifts in controls and patients post-harmonization—decreasing in controls and increasing in patients—resulting in a balanced age effect around 16% in both groups. Given the high R² for age post-harmonization and its directional shift between groups, we included age as a covariate in subsequent group comparisons.

**Table S3:** Gray Matter ROIs replication externally-enriched cohort.

| Region | HC Mean | P Mean | HC-P Diff | p-val | FDR p | Cohen's d | Cohen's d CI |
| --- | --- | --- | --- | --- | --- | --- | --- |
| TE10TE12R | 2306.7 | 2164.4 | 142.3 | <0.001 | <0.001 | 0.5 | [0.31, 0.69] |
| vldvlgR | 1353.8 | 1279.2 | 74.7 | <0.001 | <0.001 | 0.48 | [0.29, 0.67] |
| A11mR | 2038.3 | 1884.7 | 153.7 | <0.001 | <0.001 | 0.43 | [0.24, 0.62] |
| A1247oL | 1647.2 | 1563.7 | 83.5 | <0.001 | <0.001 | 0.42 | [0.23, 0.61] |
| rTthaL | 851.8 | 793.9 | 57.8 | <0.001 | <0.001 | 0.41 | [0.22, 0.6] |
| A32sgR | 1563.1 | 1488.3 | 74.8 | <0.001 | <0.001 | 0.38 | [0.19, 0.57] |
| vldvlgL | 1196.6 | 1145.4 | 51.2 | <0.001 | <0.001 | 0.35 | [0.16, 0.54] |
| A14mL | 1721.3 | 1654.9 | 66.4 | <0.001 | <0.001 | 0.33 | [0.14, 0.52] |
| rTthaR | 832.6 | 781.0 | 51.6 | <0.001 | <0.001 | 0.32 | [0.13, 0.51] |
| cHippL | 2421.6 | 2334.2 | 87.4 | 0.001 | 0.024 | 0.3 | [0.11, 0.49] |
| A32pR | 1404.5 | 1355.8 | 48.7 | 0.002 | 0.024 | 0.28 | [0.09, 0.47] |
| TE10TE12L | 2443.4 | 2351.9 | 91.5 | 0.003 | 0.024 | 0.28 | [0.09, 0.47] |
| vlaR | 899.8 | 868.3 | 31.5 | 0.005 | 0.027 | 0.26 | [0.07, 0.45] |
| mPFthaL | 927.0 | 886.6 | 40.4 | 0.007 | 0.027 | 0.25 | [0.06, 0.44] |
| A32sgL | 2281.8 | 2203.1 | 78.7 | 0.007 | 0.027 | 0.25 | [0.06, 0.44] |
| A9mR | 2954.8 | 2879.9 | 74.9 | 0.008 | 0.027 | 0.24 | [0.05, 0.43] |
| dldL | 1720.1 | 1669.1 | 51.0 | 0.009 | 0.027 | 0.24 | [0.05, 0.43] |
| dlaR | 1025.0 | 998.3 | 26.6 | 0.014 | 0.034 | 0.23 | [0.04, 0.42] |
| dlaL | 1152.7 | 1120.4 | 32.3 | 0.012 | 0.032 | 0.23 | [0.04, 0.42] |
| vlaL | 1056.2 | 1030.9 | 25.3 | 0.02 | 0.042 | 0.22 | [0.03, 0.41] |
| A1247IR | 1617.3 | 1573.9 | 43.4 | 0.021 | 0.042 | 0.21 | [0.02, 0.4] |
| PPthaR | 966.8 | 932.8 | 34.0 | 0.023 | 0.042 | 0.21 | [0.02, 0.4] |
| dIPuL | 1618.1 | 1553.1 | 65.0 | 0.053 | 0.091 | 0.18 | [-0.01, 0.37] |
| A10mL | 2846.3 | 2781.5 | 64.8 | 0.063 | 0.101 | 0.17 | [-0.02, 0.36] |
| A10mR | 2597.0 | 2548.4 | 48.6 | 0.094 | 0.141 | 0.15 | [-0.04, 0.34] |
| dIPuR | 1523.6 | 1471.0 | 52.6 | 0.135 | 0.191 | 0.14 | [-0.05, 0.33] |
| dIdR | 1341.1 | 1319.7 | 21.5 | 0.173 | 0.23 | 0.12 | [-0.07, 0.31] |
| A23cL | 2143.9 | 2112.4 | 31.5 | 0.213 | 0.256 | 0.11 | [-0.08, 0.3] |
| A13L | 2855.0 | 2801.8 | 53.2 | 0.317 | 0.331 | 0.09 | [-0.1, 0.28] |
| IPFthaR | 1355.0 | 1330.4 | 24.6 | 0.316 | 0.331 | 0.09 | [-0.1, 0.28] |
| vmPuR | 1001.4 | 995.7 | 5.7 | 0.698 | 0.698 | 0.03 | [-0.16, 0.22] |
| A23cR | 1994.9 | 2021.1 | -26.1 | 0.312 | 0.331 | -0.1 | [-0.29, 0.09] |
| A4142R | 941.5 | 973.1 | -31.6 | 0.182 | 0.23 | -0.13 | [-0.32, 0.06] |

### Multimodal Analysis of ROIs

#### DTI: Fractional Anisotropy (FA)

Given that for each modality there was some loss in our cohort due to missing neuroimaging variables, we confirmed that age remained balanced in each modality. In the case control comparison for FA in our ROIs, 32/33 regions were significant after FDR correction, indicating greater white matter integrity in HCs compared to patients. Cohen’s d values ranged from 0.29 to 0.89, with 7 regions showing large effect sizes (Cohen’s d > 0.7), indicating overall evidence for large group differences.

Notably, the five regions with the largest effect sizes were all thalamic regions, highlighting potential thalamic involvement in our cohort. We provide an overview of all values in Table S3. Fig. S2 shows the tbss tracts that run through the selected BN regions. These form 20 significant clusters (> 30 voxels).

**Fig. S2:**
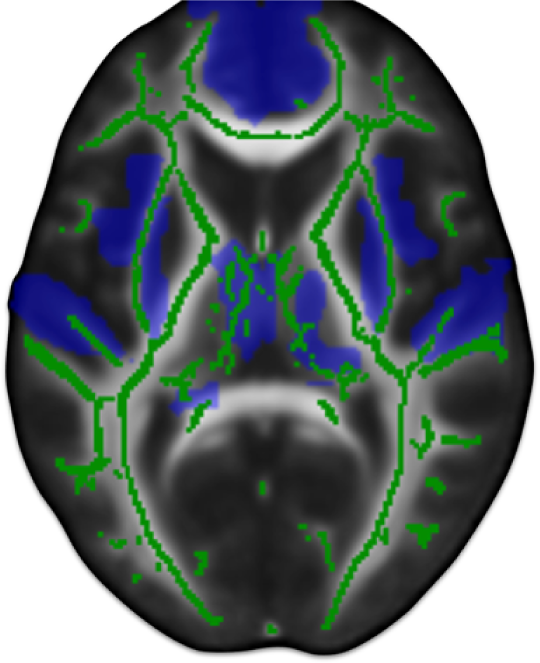
All Brainetome ROIs (blue) were converted to diffusion space and placed on the tbss skeleton (green). For the exact Regions see Figure 2 in main text.

#### Resting-state fMRI: Within-region functional connectivity

In the case control comparison for within-region functional connectivity, 27/33 regions were significant after FDR correction and Cohen’s d values ranged from 0.01 to 0.64, with 10 regions showing moderate effect sizes (Cohen’s d > 0.5), indicating substantial evidence for group differences in these regions. The regions with the highest effect sizes were the TE1.0, located in the Superior Temporal Gyrus, and two Thalamic regions. An overview of all effect sizes and significance levels is provided in Table S4.

#### Resting-state fMRI: Between-regions functional connectivity

Patients had consistently lesser values in between-regions functional connectivity than controls across all regions. Of those, 23/33 regions were significant after FDR correction and Cohen’s d values ranged from 0 to 0.56, with 5 regions showing moderate effect sizes (Cohen’s d > 0.5). Of those regions, the bilateral rostral thalamic regions and the right subgenual area 32, part of the cingulate gyrus, have demonstrated consistently high effect sizes across other modalities. An overview of all effect sizes and significance levels is provided in Table S6.

**Table S4:** FA.

| Region | HC Mean | P Mean | p-val | FDR p-val | Cohen's $d$ | Cohen's $d$ |
| --- | --- | --- | --- | --- | --- | --- |
| rTthaL | 0.352 | 0.329 | <0.001 | <0.001 | 0.83 | [0.54, 1.11] |
| PPthaR | 0.338 | 0.325 | <0.001 | <0.001 | 0.82 | [0.53, 1.11] |
| rTthaR | 0.314 | 0.293 | <0.001 | <0.001 | 0.75 | [0.46, 1.03] |
| IPFthaR | 0.333 | 0.323 | <0.001 | <0.001 | 0.73 | [0.44, 1.01] |
| mPFthaL | 0.337 | 0.325 | <0.001 | <0.001 | 0.72 | [0.44, 1.01] |
| vldvlgL | 0.454 | 0.431 | <0.001 | <0.001 | 0.72 | [0.44, 1.00] |
| dlaR | 0.33 | 0.318 | <0.001 | <0.001 | 0.71 | [0.43, 0.99] |
| dIPuR | 0.444 | 0.43 | <0.001 | <0.001 | 0.67 | [0.39, 0.95] |
| A32sgL | 0.523 | 0.505 | <0.001 | <0.001 | 0.64 | [0.36, 0.93] |
| dIPuL | 0.492 | 0.478 | <0.001 | <0.001 | 0.64 | [0.36, 0.92] |
| cHippL | 0.451 | 0.43 | <0.001 | <0.001 | 0.6 | [0.32, 0.88] |
| dIdR | 0.375 | 0.359 | <0.001 | <0.001 | 0.59 | [0.31, 0.87] |
| vldvlgR | 0.393 | 0.38 | <0.001 | <0.001 | 0.56 | [0.28, 0.84] |
| A23cR | 0.516 | 0.496 | <0.001 | 0.002 | 0.56 | [0.28, 0.84] |
| IPFthaL | 0.408 | 0.399 | 0.001 | 0.002 | 0.56 | [0.28, 0.84] |
| A32sgR | 0.451 | 0.43 | <0.001 | <0.001 | 0.55 | [0.27, 0.83] |
| A10mR | 0.443 | 0.414 | <0.001 | <0.001 | 0.55 | [0.27, 0.83] |
| A32pR | 0.548 | 0.523 | <0.001 | <0.001 | 0.54 | [0.26, 0.82] |
| A23cL | 0.463 | 0.445 | <0.001 | <0.001 | 0.54 | [0.26, 0.82] |
| A10mL | 0.525 | 0.501 | <0.001 | <0.001 | 0.53 | [0.25, 0.81] |
| A9mR | 0.521 | 0.5 | <0.001 | <0.001 | 0.52 | [0.24, 0.80] |
| TE10TE12L | 0.369 | 0.357 | <0.001 | <0.001 | 0.5 | [0.22, 0.77] |
| vmPuR | 0.508 | 0.494 | 0.002 | 0.002 | 0.49 | [0.21, 0.77] |
| dlaL | 0.324 | 0.314 | 0.001 | 0.002 | 0.49 | [0.21, 0.77] |
| vlaR | 0.345 | 0.335 | 0.001 | 0.002 | 0.48 | [0.20, 0.76] |
| A1247IR | 0.416 | 0.401 | <0.001 | <0.001 | 0.47 | [0.19, 0.75] |
| dIdL | 0.386 | 0.373 | 0.002 | 0.002 | 0.45 | [0.18, 0.73] |
| vlaL | 0.329 | 0.32 | 0.006 | 0.008 | 0.44 | [0.16, 0.72] |
| A4142R | 0.417 | 0.405 | 0.012 | 0.013 | 0.42 | [0.14, 0.69] |
| A13L | 0.31 | 0.302 | 0.011 | 0.013 | 0.33 | [0.06, 0.61] |
| TE10TE12R | 0.364 | 0.355 | 0.015 | 0.016 | 0.3 | [0.03, 0.58] |
| A11mR | 0.253 | 0.242 | 0.166 | 0.166 | 0.29 | [0.01, 0.57] |
| A1247oL | 0.309 | 0.298 | 0.038 | 0.040 | 0.29 | [0.01, 0.57] |
| A14mL | 0.44 | 0.431 | 0.099 | 0.102 | 0.24 | [-0.04, 0.51] |

**Table S5:** WR-fC.

| Region | HC Mean | P Mean | HC-P Diff | p-val | FDR p-val | Cohen's d | Cohen's d CI |
| --- | --- | --- | --- | --- | --- | --- | --- |
| TE10TE12R | 0.295 | 0.227 | -0.068 | <0.0001 | <0.0001 | 0.64 | (0.37, 0.9) |
| mPFthaL | 0.305 | 0.241 | -0.064 | <0.0001 | 0.0002 | 0.62 | (0.35, 0.88) |
| IPFthaR | 0.242 | 0.186 | -0.055 | <0.0001 | 0.0002 | 0.59 | (0.32, 0.85) |
| A32sgR | 0.427 | 0.346 | -0.08 | <0.0001 | 0.0004 | 0.57 | (0.31, 0.83) |
| A32pR | 0.424 | 0.349 | -0.075 | 0.0002 | 0.0007 | 0.55 | (0.29, 0.81) |
| rTthaL | 0.205 | 0.169 | -0.035 | 0.0002 | 0.0006 | 0.55 | (0.28, 0.81) |
| dIdL | 0.318 | 0.264 | -0.054 | <0.0001 | 0.0003 | 0.55 | (0.28, 0.81) |
| A23cR | 0.218 | 0.176 | -0.042 | 0.0002 | 0.0007 | 0.54 | (0.28, 0.81) |
| rTthaR | 0.243 | 0.199 | -0.044 | 0.0002 | 0.0006 | 0.53 | (0.27, 0.8) |
| dIaR | 0.427 | 0.359 | -0.068 | 0.0005 | 0.0012 | 0.52 | (0.26, 0.78) |
| A32sgL | 0.386 | 0.318 | -0.068 | 0.0003 | 0.0009 | 0.49 | (0.23, 0.76) |
| A4142R | 0.449 | 0.369 | -0.08 | <0.0001 | 0.0002 | 0.49 | (0.23, 0.75) |
| vIaL | 0.33 | 0.285 | -0.045 | 0.0011 | 0.0025 | 0.47 | (0.21, 0.73) |
| TE10TE12L | 0.29 | 0.236 | -0.054 | 0.0002 | 0.0006 | 0.46 | (0.2, 0.72) |
| vIaR | 0.293 | 0.254 | -0.039 | 0.0028 | 0.0055 | 0.44 | (0.18, 0.7) |
| cHippL | 0.142 | 0.123 | -0.019 | 0.0167 | 0.0262 | 0.42 | (0.16, 0.68) |
| A23cL | 0.182 | 0.158 | -0.025 | 0.0025 | 0.0051 | 0.42 | (0.16, 0.68) |
| A14mL | 0.453 | 0.392 | -0.061 | 0.0082 | 0.0138 | 0.42 | (0.16, 0.68) |
| A9mR | 0.259 | 0.225 | -0.034 | 0.0059 | 0.0109 | 0.41 | (0.15, 0.67) |
| vIdvlgR | 0.277 | 0.238 | -0.039 | 0.0021 | 0.0046 | 0.4 | (0.14, 0.66) |
| A13L | 0.101 | 0.086 | -0.015 | 0.0084 | 0.0138 | 0.39 | (0.13, 0.65) |
| dIdR | 0.305 | 0.269 | -0.036 | 0.0202 | 0.0303 | 0.34 | (0.08, 0.6) |
| PPthaR | 0.177 | 0.16 | -0.017 | 0.0683 | 0.0777 | 0.29 | (0.03, 0.55) |
| dIaL | 0.373 | 0.338 | -0.035 | 0.0266 | 0.0366 | 0.28 | (0.02, 0.54) |
| A1247IR | 0.298 | 0.269 | -0.029 | 0.0355 | 0.0468 | 0.27 | (0.01, 0.53) |
| vIdvlgL | 0.27 | 0.248 | -0.022 | 0.0232 | 0.0332 | 0.26 | (-0, 0.52) |
| A10mL | 0.285 | 0.262 | -0.023 | 0.0379 | 0.0481 | 0.22 | (-0.04, 0.47) |
| dIPuL | 0.295 | 0.269 | -0.027 | 0.0402 | 0.0492 | 0.21 | (-0.05, 0.46) |
| dIPuR | 0.304 | 0.277 | -0.027 | 0.0644 | 0.0759 | 0.2 | (-0.06, 0.46) |
| A10mR | 0.317 | 0.295 | -0.022 | 0.2197 | 0.2339 | 0.17 | (-0.09, 0.43) |
| vmPuR | 0.343 | 0.322 | -0.021 | 0.1734 | 0.1907 | 0.16 | (-0.1, 0.42) |
| A1247oL | 0.198 | 0.196 | -0.003 | 0.3354 | 0.3459 | 0.04 | (-0.22, 0.3) |
| A11mR | 0.192 | 0.192 | 0.001 | 0.3768 | 0.3768 | -0.01 | (-0.27, 0.25) |

**Table S6:** BR-fC.

| Region | HC Mean | P Mean | HC-P Diff | p-val | FDR p-val | Cohen's d | Cohen's d CI |
| --- | --- | --- | --- | --- | --- | --- | --- |
| vldvlgL | 0.189 | 0.131 | 0.058 | <0.001 | <0.001 | 0.56 | (0.30, 0.83) |
| A23cR | 0.186 | 0.127 | 0.058 | <0.001 | 0.002 | 0.54 | (0.27, 0.80) |
| rTthaR | 0.209 | 0.152 | 0.058 | <0.001 | 0.002 | 0.52 | (0.25, 0.78) |
| rTthaL | 0.202 | 0.149 | 0.053 | 0.001 | 0.003 | 0.51 | (0.25, 0.77) |
| dlaR | 0.207 | 0.155 | 0.053 | <0.001 | 0.002 | 0.5 | (0.24, 0.76) |
| A32sgR | 0.273 | 0.222 | 0.051 | 0.001 | 0.003 | 0.48 | (0.22, 0.75) |
| A23cL | 0.176 | 0.125 | 0.05 | 0.001 | 0.002 | 0.48 | (0.22, 0.74) |
| vldvlgR | 0.192 | 0.143 | 0.049 | <0.001 | 0.002 | 0.46 | (0.20, 0.73) |
| mPFthaL | 0.185 | 0.13 | 0.055 | 0.002 | 0.006 | 0.46 | (0.19, 0.72) |
| vlaR | 0.201 | 0.154 | 0.047 | 0.003 | 0.006 | 0.45 | (0.19, 0.71) |
| TE10TE12R | 0.191 | 0.145 | 0.046 | <0.001 | 0.002 | 0.43 | (0.16, 0.69) |
| vmPuR | 0.155 | 0.113 | 0.042 | 0.001 | 0.004 | 0.4 | (0.14, 0.66) |
| dlaL | 0.194 | 0.152 | 0.042 | 0.002 | 0.005 | 0.4 | (0.14, 0.66) |
| IPFthaR | 0.166 | 0.121 | 0.045 | 0.007 | 0.015 | 0.39 | (0.13, 0.65) |
| PPthaR | 0.105 | 0.068 | 0.037 | 0.009 | 0.017 | 0.35 | (0.09, 0.61) |
| A32pR | 0.257 | 0.219 | 0.038 | 0.034 | 0.048 | 0.35 | (0.09, 0.61) |
| dldL | 0.181 | 0.146 | 0.035 | 0.003 | 0.007 | 0.33 | (0.07, 0.59) |
| dldR | 0.157 | 0.121 | 0.036 | 0.007 | 0.015 | 0.32 | (0.06, 0.58) |
| A32sgL | 0.246 | 0.211 | 0.035 | 0.018 | 0.029 | 0.32 | (0.06, 0.58) |
| A10mR | 0.152 | 0.123 | 0.029 | 0.026 | 0.04 | 0.29 | (0.03, 0.55) |
| A13L | 0.13 | 0.1 | 0.03 | 0.015 | 0.025 | 0.28 | (0.02, 0.54) |
| A9mR | 0.181 | 0.152 | 0.029 | 0.125 | 0.158 | 0.26 | (0.00, 0.52) |
| A10mL | 0.175 | 0.15 | 0.025 | 0.058 | 0.077 | 0.23 | (-0.03, 0.49) |
| A1247IR | 0.131 | 0.106 | 0.024 | 0.206 | 0.243 | 0.22 | (-0.03, 0.48) |
| dIPuL | 0.125 | 0.102 | 0.024 | 0.009 | 0.017 | 0.22 | (-0.04, 0.48) |
| dIPuR | 0.137 | 0.115 | 0.023 | 0.02 | 0.032 | 0.21 | (-0.04, 0.47) |
| TE10TE12L | 0.173 | 0.15 | 0.023 | 0.052 | 0.072 | 0.21 | (-0.05, 0.47) |
| A14mL | 0.154 | 0.135 | 0.019 | 0.164 | 0.2 | 0.18 | (-0.08, 0.44) |
| vlaL | 0.201 | 0.184 | 0.017 | 0.294 | 0.324 | 0.17 | (-0.09, 0.43) |
| A4142R | 0.143 | 0.129 | 0.013 | 0.235 | 0.267 | 0.12 | (-0.14, 0.38) |
| A1247oL | 0.092 | 0.097 | -0.006 | 0.458 | 0.488 | -0.05 | (-0.31, 0.21) |
| cHippL | 0.051 | 0.049 | 0.003 | 0.917 | 0.917 | 0.03 | (-0.23, 0.28) |
| A11mR | 0.044 | 0.044 | 0.0 | 0.633 | 0.653 | 0.0 | (-0.25, 0.26) |

#### Source-localized EEG

We analyzed source-localized EEG amplitude across the 17 cortical regions in our ROIs. In the case-control comparison for the eyes-closed condition in the low-frequency spectrum (1–12 Hz), only the right medial area 10 in the superior frontal gyrus was significant after FDR correction (p < 0.0001; Cohen’s d = 0.54). The TE1 region in the right superior temporal gyrus showed a trend but did not survive FDR correction. There were no significant results in the high-frequency spectrum (12–70 Hz) for either the eyes-closed or eyes-open conditions.

To examine the low-frequency spectrum in more detail, we further separated it into individual bands: Delta (1–4 Hz), Theta (4–8 Hz), and Alpha (8–12 Hz). In the case-control comparison for the eyes-closed condition, all single bands within the right medial area 10 were significant (Delta, p = 0.042; Theta, p = 0.0007; Alpha, p = 0.021). No other regions reached significance for any single band, across both eyes-closed and eyes-open conditions.

To explore broader patterns, we calculated the mean values for each single band across all 17 cortical ROIs. Significant findings included the Alpha band in the eyes-open condition (p = 0.025, Cohen’s d = 0.41), the Theta band in the eyes-closed condition (p = 0.014, Cohen’s d = 0.41), and the Theta band in the eyes-open condition (p = 0.014, Cohen’s d = 0.45), all after FDR correction. Results are visualised in Figure S3.

**Fig. S3:**
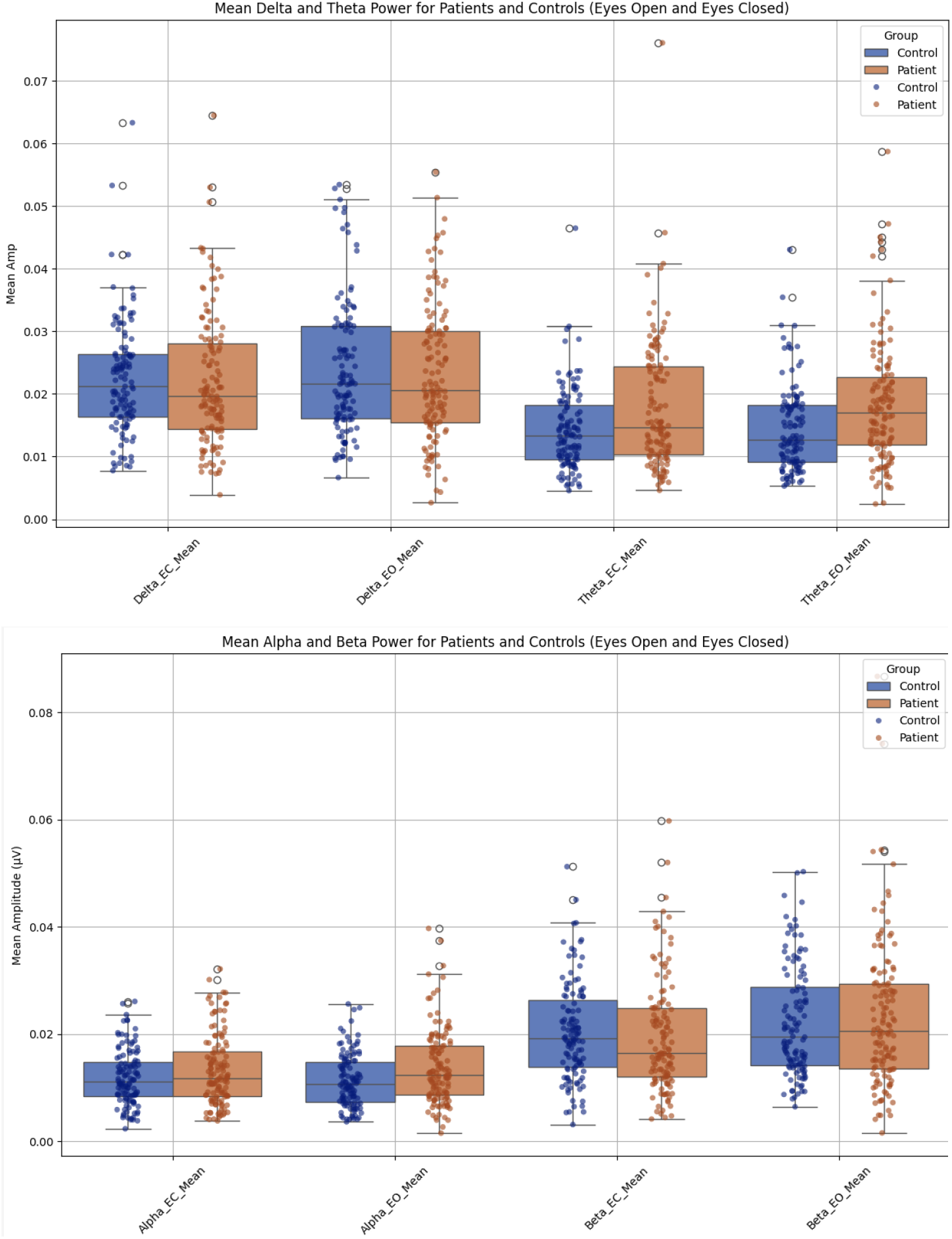
Mean EEG bands:

### Part B: Multimodal Clustering

We initially included all available modalities, but decided to exclude EEG from the final clustering for several data-driven reasons. First, EEG was the only modality that did not show significant differences between clusters, indicating its limited ability to meaningfully separate them. Additionally, it contributed minimally to the overall cluster structure, as it did not range among the top five correlations for the two PaCMAP projections. This further aligned with our case-control analyses against healthy controls, where EEG, in stark contrast to other modalities, produced only one significant result.

Importantly, removing EEG increased our sample size by 12 patients (13%), from 92 to 104 patients, thereby enhancing the robustness of our findings.

**Fig. S4:**
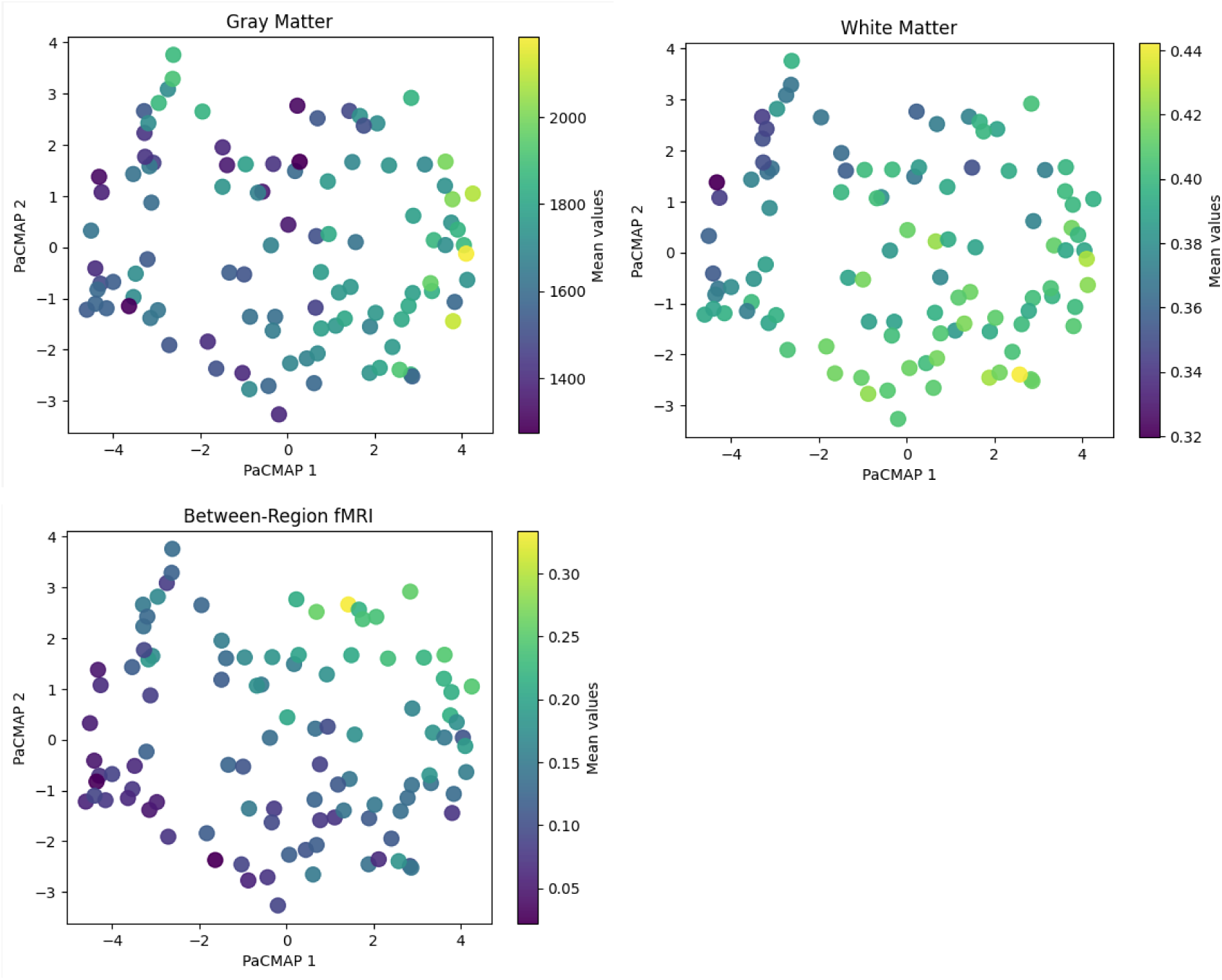
2d visualisation of PACMAP dimensionality reduction outcome.

### Cluster Characterisation

The distribution of patients across clusters was as follows: 19, 16, 30, 14, and 25 in Clusters 1 through 5, respectively. The youngest mean age was observed in Cluster 4 (34.9 years) and the oldest in Cluster 5 (39.1 years), with no significant age differences between clusters (lowest p-value = 0.815). Importantly, despite age being a well-known factor influencing volumetric brain loss, our oldest cluster (Cluster 5) showed the highest gray matter and the second-highest white matter volumes. This suggests that age was not a primary driver of the cluster structure, an effect likely achieved by removing age-related variance from brain measures for the clustering algorithm.

There were no statistically significant differences in educational attainment across clusters; each cluster had between 7 and 12 patients who had completed the highest educational level (Abitur, the university-qualifying diploma). Other clinical and health-related variables, including illness duration (ranging from 7.8 to 12.1 years), BMI (25.5 to 28.7), and chlorpromazine (CPZ) equivalent dosage (283 - 313), also showed no significant differences between clusters. No association was found between cluster membership and gender distribution (Chi-square p = 0.314). Finally, there was no difference in diagnosis between clusters, as assessed in the ratios of SZ to SZA (Chi-square p = 0.848), indicating that grouping by diagnosis can not replace the biology-based clusters.

Overall these findings reinforce that the clustering approach effectively distinguished patients based on neuroimaging features rather than sociodemographic or clinical confounders.

In terms of the Clinical Global Impression severity scale (CGI-S) our overall average lies at 4.2, with 4 being defined as “moderately ill”. No patient received a rating of 7, describing the highest severity. However, Cluster 2 had by far the most patients (26.7%) rated with 6 (“severely ill”) and no patient below a rating of 4. In marked contrast, 36% of patients in Clusters 4 and 29% in Cluster 5 have been rated with a CGI-S score of 3 (“mildly ill”).

Consistent differences emerged in PANSS total scores between Clusters 3 vs. 2 and 5 vs. 2, primarily driven by significant variations in the PANSS negative and general symptom subscales. Cluster 2 showed a markedly higher PANSS total score (63.9 ± 16.4) compared to Cluster 3 (47.7 ± 14.7) and was the only cluster with a score exceeding the threshold proposed by Leucht et al. for a “mildly ill” classification (PANSS >58). This difference of approximately 29% aligns with the level of change that Leucht et al. associate with six weeks of “minimal improvement” following treatment intervention, highlighting a meaningful clinical distinction between clusters in a cohort of generally stable patients.

**Table S7:** Sociodemographic Cluster Characterisation.

| Cluster | Patient count | Age | Sex (Female) | Education* | BMI | CPZ-e q | Disease duration (years) |
| --- | --- | --- | --- | --- | --- | --- | --- |
| 1 | 19 | 37.1 ($\pm 15$) | 37% | 12 | 25.5 | 283 | 12.1 |
| 2 | 16 | 39.0 ($\pm 13.5$) | 6% | 10 | 28.7 | 305 | 7.8 |
| 3 | 30 | 36.6<br>(±9.3) | 33% | 8 | 27.2 | 304 | 10.6 |
| 4 | 14 | 34.9<br>(±9.5) | 21% | 7 | 27.6 | 343 | 11.1 |
| 5 | 25 | 39.1<br>(10.5) | 12% | 7 | 27.9 | 314 | 11.6 |

\*Number of patients with the highest level of German highschool accomplishment ("Abitur")

**Table S8:** Clinical Cluster Characterisation.

| Cluster | GAF | PANSS<br>negative | PANSS<br>general | PANSS<br>positive | BACS<br>score | BRS | Loneliness | CTS |
| --- | --- | --- | --- | --- | --- | --- | --- | --- |
| 1 | 55<br>(±11.8) | 12.9<br>(±5.6) | 25.9<br>(±7.5) | 11.8<br>(±4.7) | 0.1<br>(±0.8) | 3.0<br>(±0.9) | 6.5<br>(±2.0) | 9.7<br>(±4.1) |
| 2 | 44.3<br>(±5.8) | 16.6<br>(±5.5) | 33.3<br>(±8.4) | 14.1<br>(±5.1) | -0.7<br>(±1.4) | 2.3<br>(±0.7) | 8.2<br>(±3.2) | 9.8<br>(±3.3) |
| 3 | 59<br>(±12.6) | 10.8<br>(±4.3) | 25.3<br>(±7.6) | 11.6<br>(±5.0) | 0.3<br>(±0.8) | 2.7<br>(±0.8) | 7.3<br>(±2.6) | 9.4<br>(±3.8) |
| 4 | 56<br>(± 10.9) | 12.7<br>(±4.0) | 28.9<br>(±6.3) | 10.8<br>(±3.0) | 0.1<br>(±1.1) | 2.8<br>(±0.6) | 7.5<br>(±1.8) | 9.7<br>(±4.0) |
| 5 | 56.8<br>(± 13.8) | 12.5<br>(±5.6) | 26.4<br>(±7.6) | 11.5<br>(±3.2) | 0.0<br>(±0.9) | 3.2<br>(±0.7) | 7.3<br>(±2.0) | 9.0<br>(±3.8) |

**Fig. S5:**
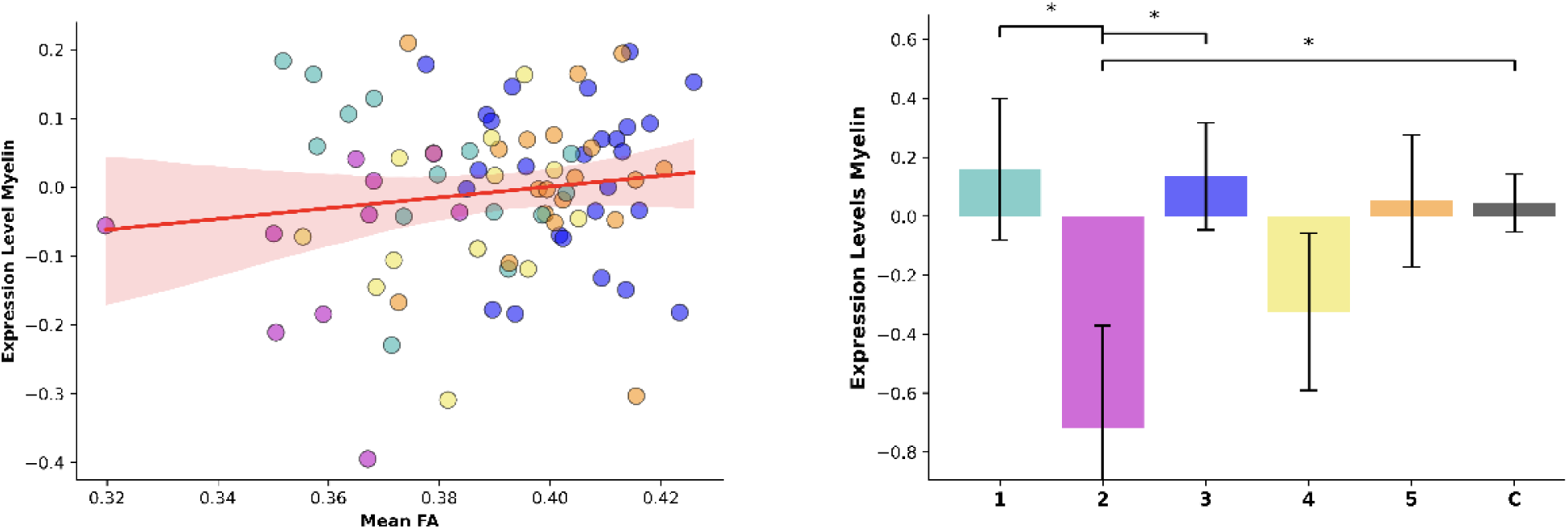
Myelin Pathway Expression. Left: Correlation with mean FA. Right: Group Comparison. FA=Fractional Anisotropy.

### Cluster validity and robustness

To ensure the robustness and validity of our clustering results, we conducted five key assessments: cluster characterization, statistical cluster validation, cluster stability testing across bootstrap subsampling, stability of brain-symptom associations across bootstraps and comparison of PaCMAP-based clusters with clusters based on the Uniform Manifold Approximation and Projection (UMAP) method. Below, we detail each step.

First, we characterised the clusters in terms of sociodemographic variables, ensuring that potentially confounding factors are not unbalanced between the clusters (see Table S6). Furthermore, while unsupervised clustering lacks a formal ground truth, we assessed the "ground meaning" by evaluating whether the clusters aligned with plausible patterns in clinical and biological variables. We calculated t-statistics and p-values to compare each cluster against the other, applying FDR correction for each variable separately (see outcome descriptions in results in main text).

Second, a fundamental question in clustering is whether identified groups are “really there, as opposed to being artifacts of the natural sampling variation” (72). Theoretically, this could happen even with moderate to strong cluster validation indices and meaningful clinical and biological patterns. To address this problem, we applied a Monte Carlo-based approach adapted from Dinga et al (73). Specifically, we compared our cluster validation metrics against null distributions generated from synthetic datasets.

These synthetic datasets were sampled from a multivariate Gaussian distribution, matching the mean vector and covariance structure of the PaCMAP projections in our data. For each of 10 000 iterations, we generated a synthetic dataset, performed Gaussian Mixture Model (GMM) clustering, and calculated our cluster validation indices for the synthetic clusters. The observed indices from our actual data were compared to these null distributions, yielding p-values that quantify the likelihood of obtaining such scores when in fact no cluster structure is present. p-values below 0.05 indicate that the observed clustering structure is unlikely to arise purely by chance. The results are visualised in Figure S6. To understand how sensitive this test against null distribution is in our data, we repeated the experiment with a 12-cluster structure, the highest number of clusters with a still acceptable silhouette score (0.34). This resulted in p-values above the significance threshold (0.16 for the Silhouette score, 0.28 for Davies-Boudin), indicating that some scenarios in our cohort result in acceptable but not significant validation metrics and providing further evidence that our 5 clusters reflect a meaningful structure rather than artifacts of sampling variability.

Third, to test cluster stability, we performed 10 000 bootstrap runs, each involving 85–95% of the sample (89–98 unique patients) randomly selected. For each run, we reapplied the clustering algorithm, starting with a fresh PaCMAP dimensionality reduction. We calculated mean silhouette scores for the 5 cluster solution and mean silhouette scores for the optimal cluster (based on the highest silhouette score) in each run. The mean silhouette score across all runs served as a marker of the pipeline’s ability to produce well-separated clusters.

Fourth, to evaluate the robustness of brain-clinical relationships, we created multimodal composite scores by correlating clinical variables (e.g. GAF) with ROIs across modalities. For each modality, ROIs with pearson correlations ≥ 0.2 were z-standardized, and their mean values were combined to form a composite score for the corresponding clinical variable. Across 10 000 bootstrap runs, we then compared the difference between the clusters with the highest and lowest values for key clinical variables and their corresponding mean multimodal brain-based composite scores (e.g. GAF and brain-based GAF composite score) (see Figure S7).

Finally, since PaCMAP has not yet been applied in neuroimaging, we compared PaCMAP-based clusters with UMAP-based clusters, allowing us to evaluate the consistency of results across different dimensionality reduction methods.

**Fig. S6:**
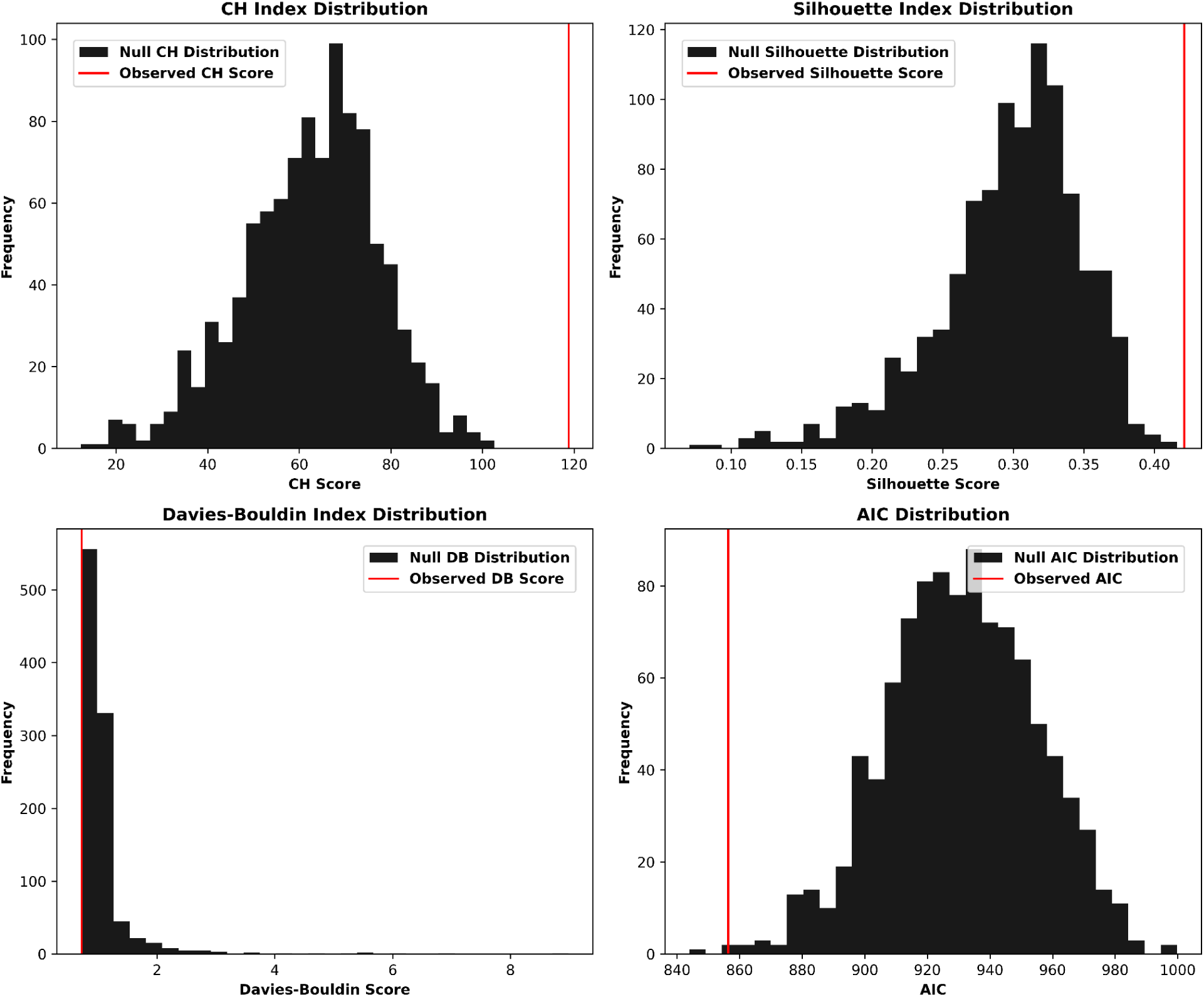
Cluster validation metrics observed in our cohort (red) against null distributions of validation metrics observed in synthetic datasets. AIC=Akaike Information Criterion. *Note: Lower DB and AIC indicate better cluster separation*.

**Fig. S7:**
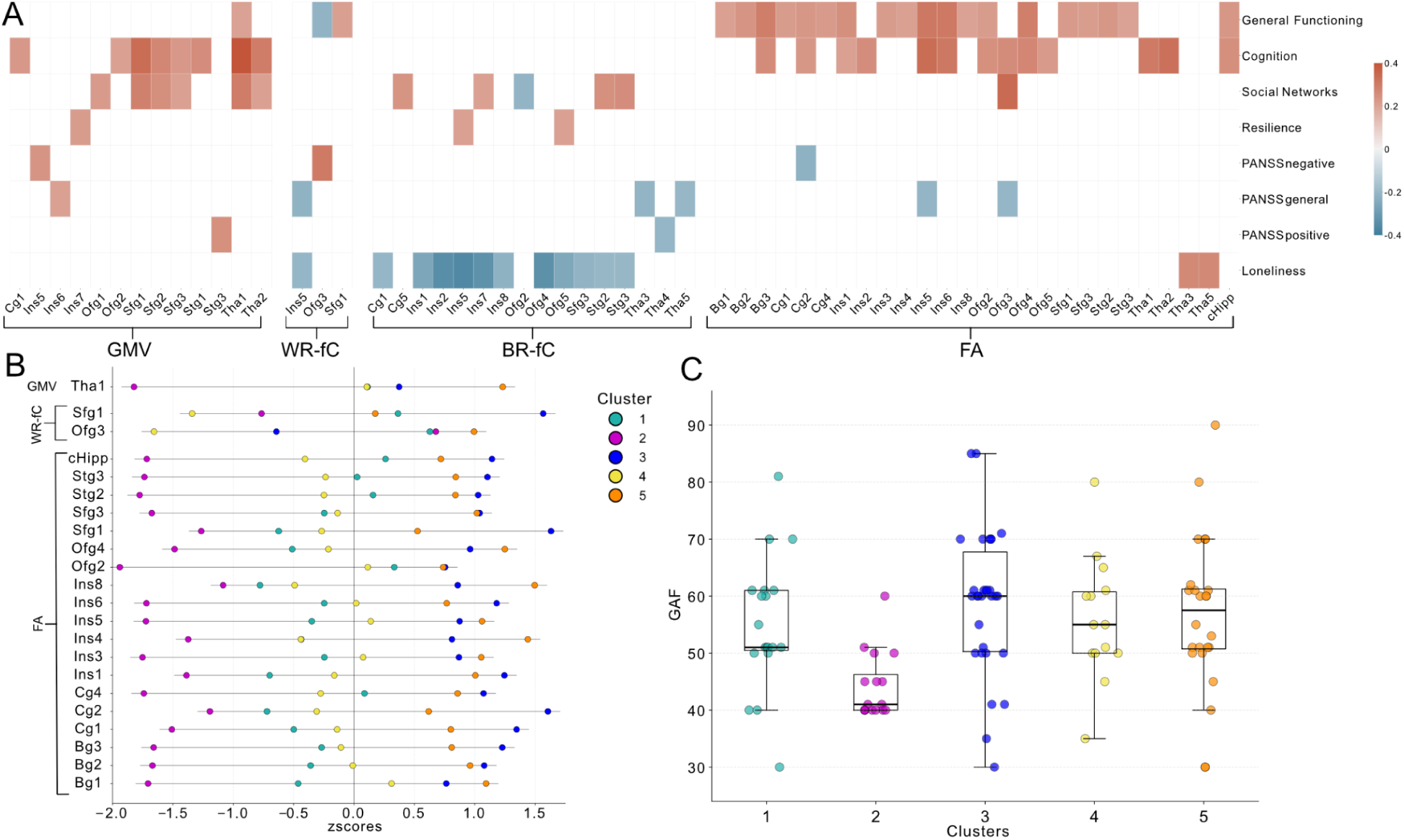
Brain-Symptom associations. **A:** All correlations between neuroimaging outcomes and clinical variables exceeding the threshold of a weak correlation (r ≥ 0.2, p < 0.05). **B:** Distribution by cluster of all z-standardised regions significantly correlating with the GAF. **C:** z-standardised distribution of the GAF score. GMV=Gray Matter Volume. BR-fC=Between-Region functional connectivity. WR-fC=Within-Region functional connectivity (WR-fC), FA = Fractional Anisotropy. General Functioning=GAF=Global Assessment of Functioning. Resilience=BRS=Brief Resilience Scale. PANSS=Positive and Negative Syndrome Scale. Sociat Networks = Lubben Social Networks, Friends subscale.

**Fig. S8:**
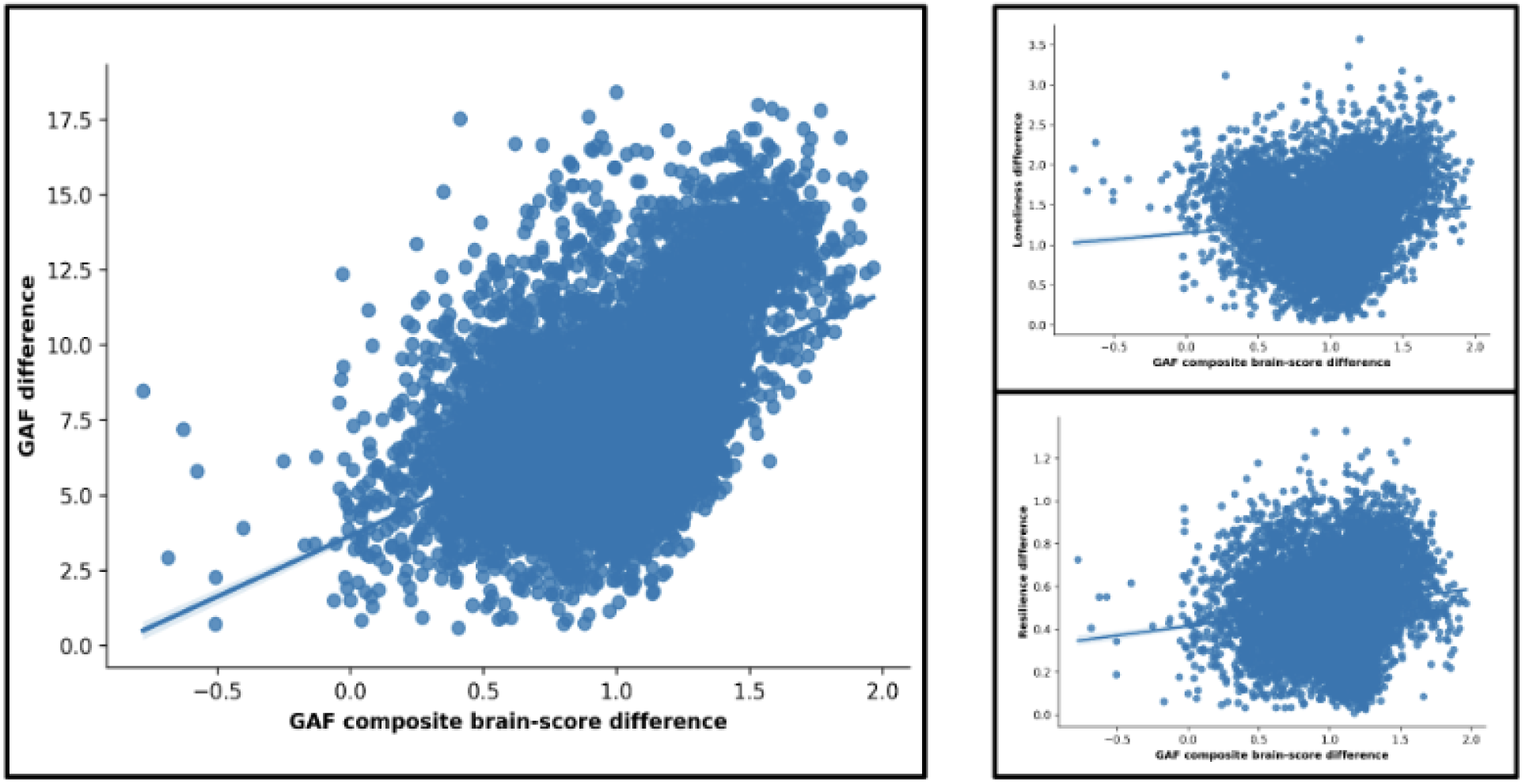
**Left**: Across bootstrap runs, the GAF difference (y axis) between extreme clusters in GAF (highest vs. lowest GAF) and the difference in its neuroscore (x axis) correlate significantly (r=0.47, p = <.001). **Right**: To assess how specific this relationship is, we calculated the correlation between the GAF-specific neuroscore and unrelated clinical variables such as loneliness (right top) (r=0.11, p <.001) and resilience (right bottom) (0.14, p <.001). GAF=Global Assessment of Functioning.

### Comparison against UMAP- based GMM clustering

After confirming the robustness of our PaCMAP-based GMM clustering through validation metrics, visual inspection, meaningful correlations between neuroimaging and clinical variables, and bootstrap analysis, we conducted a final robustness assessment by comparing it to UMAP-based GMM clustering. UMAP, as a widely adopted nonlinear dimensionality reduction technique, served as a critical benchmark against which to evaluate our findings.

Applying the same analytical pipeline, we used UMAP to create a 2D projection from the 132 standardized multimodal variables, followed by GMM clustering. The validation metrics for the UMAP-based clustering indicated moderately good performance (e.g. a Silhouette Score of 0.41) and an acceptable visual inspection, although the separation was less distinct than between PaCMAP based clusters and more patients had a lower probability of being assigned correctly (Figure S8). Nevertheless, clustering comparison metrics revealed moderate to strong agreement between the two approaches, with Cohen’s Kappa values for specific cluster pairs ranging from 0.64 to 0.75. PaCMAP provided a finer-grained segmentation by splitting the most clinically burdened cluster into two distinct subgroups and both methods converged on a comparable underlying structure within the data (Figure S9).

**Fig. S9:**
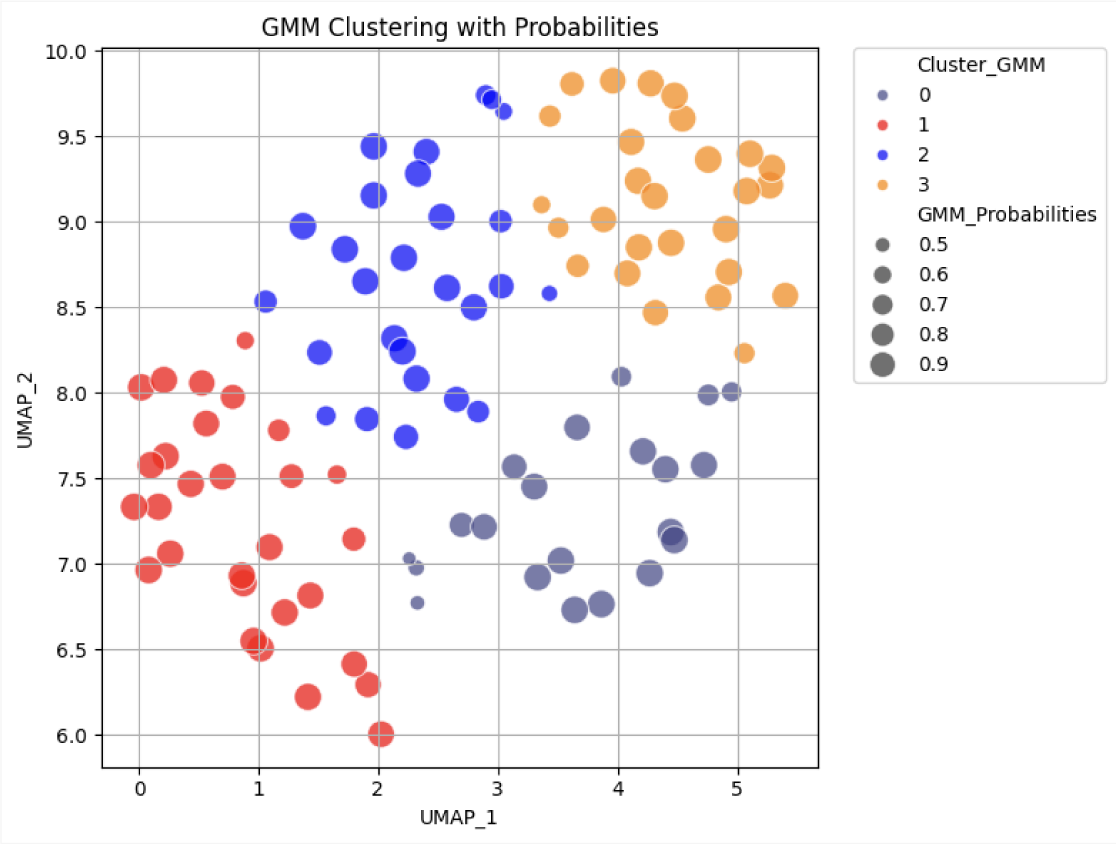
UMAP clustering. GMM=Gaussian Mixture Models. UMAP=UMAP=Uniform Manifold Approximation and Projection.

**Fig. S10:**
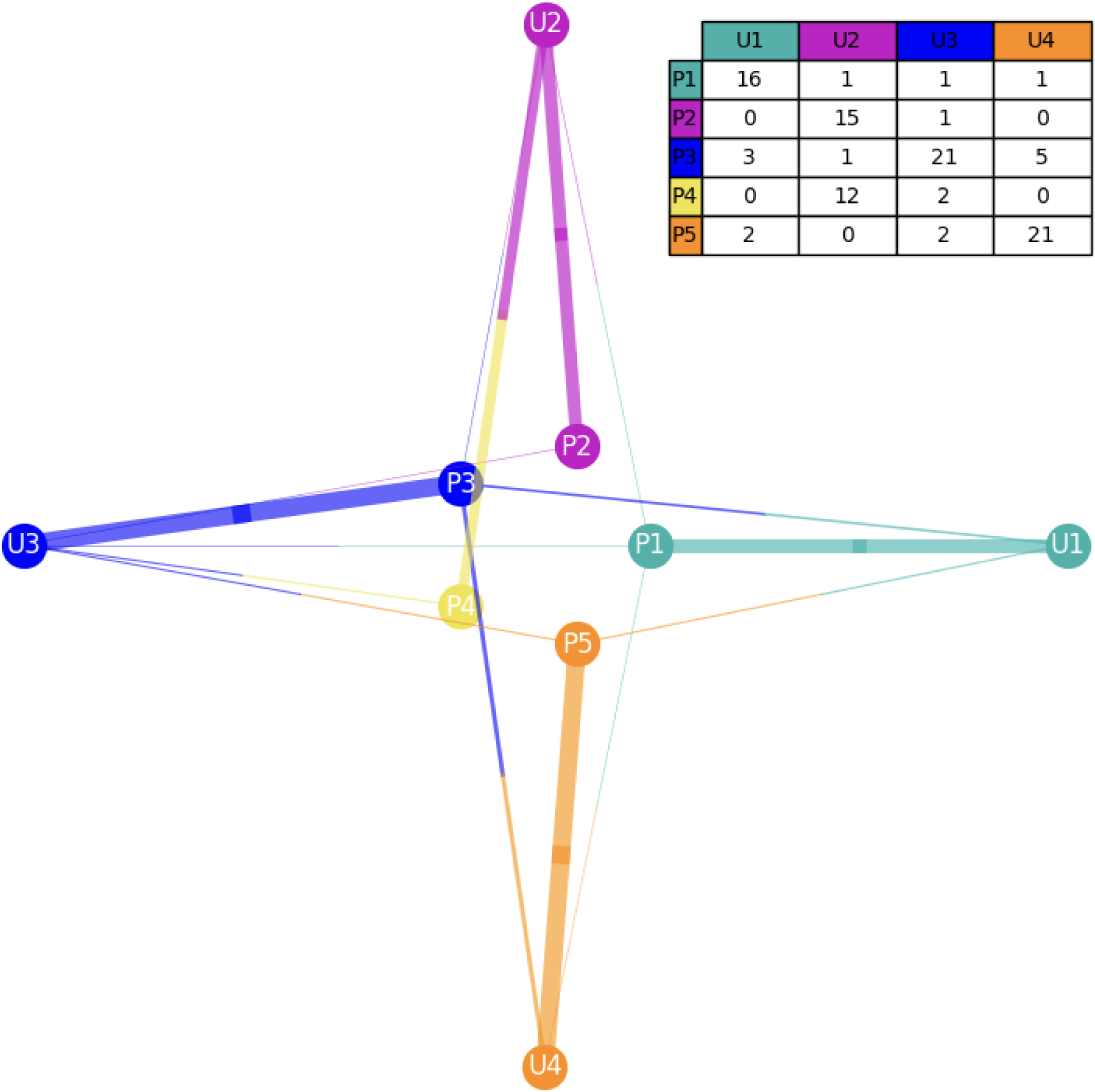
Comparison of Cluster belonging between PacMAP (Center) and UMAP (Surrounding). P = PacMAP Cluster. U = UMAP Cluster. PacMAP=Pairwise Controlled Manifold Approximation and Projection. UMAP=Uniform Manifold Approximation and Projection.

The cluster sizes after UMAP were balanced with 21 patients in Cluster 1, 29 in Cluster 2, 27 in 3 and 27 in 4. Sociodemographic factors, including age, BMI, and disease duration, remained nonsignificant across clusters, further confirming that our clustering outcomes were not confounded by these variables.

Key neuroimaging-clinical relationships were preserved across the two clustering methods, providing consistent insights into the interplay between brain structure, connectivity, and clinical variables. Cluster 2 was characterized by overall lower neuroimaging values across modalities and clinical scores indicative of greater symptom severity. Specifically, in UMAP-based clustering, Cluster 2 exhibited significantly reduced FA values compared to Clusters 3 and 4 (p = <.001), mirroring findings from PaCMAP based clustering. Similarly, the cluster with highest GMV was consistently associated with higher resilience scores, with Cluster 4 in UMAP outperforming Cluster 2 (p = <0.001). Functional connectivity also followed comparable patterns: Cluster 1, which had the highest between-region connectivity, showed significantly lower loneliness scores than other clusters, particularly Cluster 2.

UMAP-based clustering also replicated the relationships between neuroimaging patterns and polygenic risk scores, with Cluster 4, showing highest overall GMV and significantly higher PRS for intracranial volume. All other PRS scores remained non-significant.

PaCMAP-based clustering demonstrated a more fine-grained segmentation, splitting the most clinically burdened cluster into two subgroups (U2 in P2 and P4, see Figure).

Accordingly, some differences in clinical scores were more pronounced in PaCMAP-based clustering. For example, the difference in GAF scores between the highest and lowest clusters was 14.6, compared to 9.6 in UMAP-based clustering. This finer segmentation might reflect PaCMAP’s ability to identify more subtle variations within the data.

**Fig. S11:**
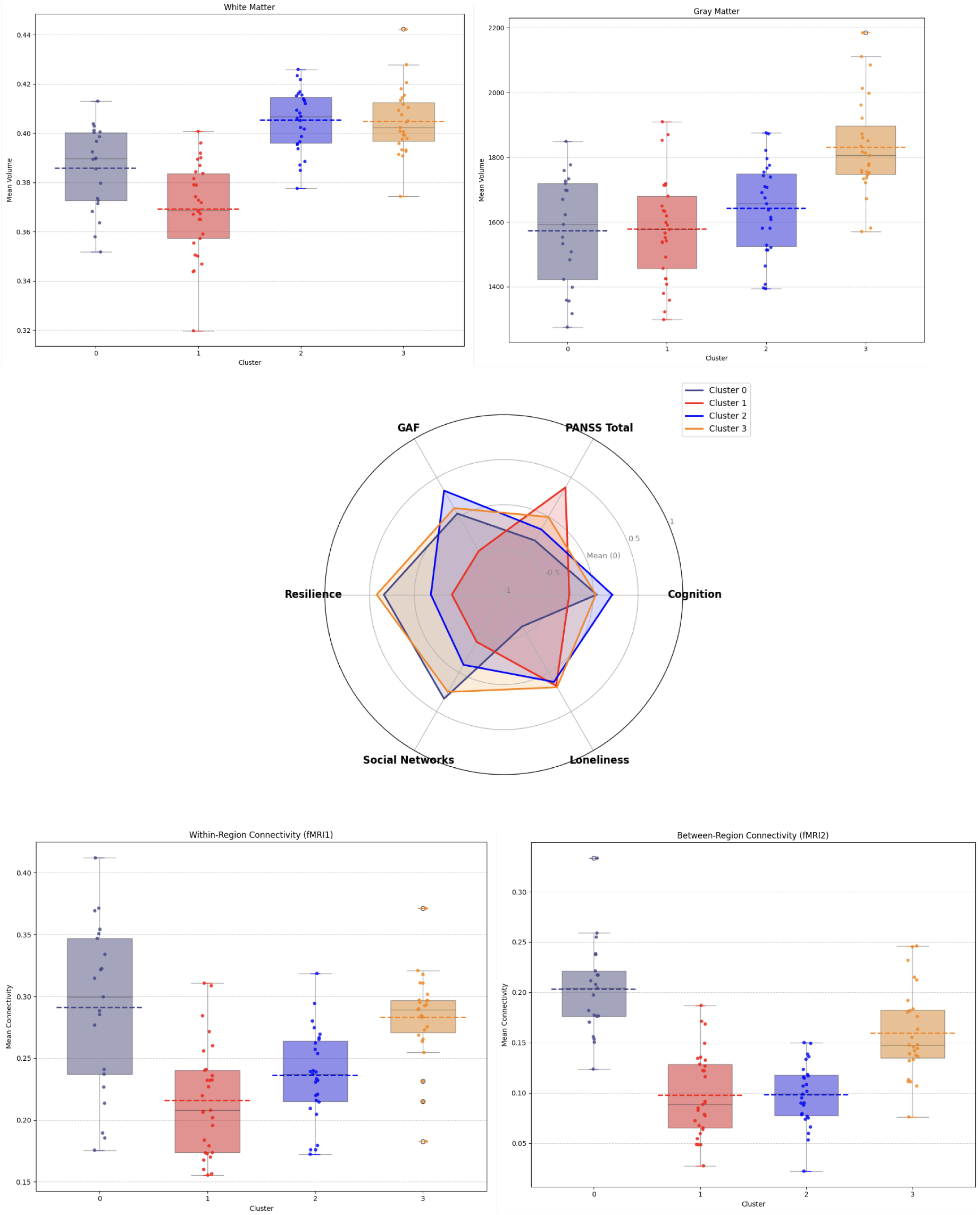
UMAP-based cluster characterisation. UMAP=Uniform Manifold Approximation and Projection. GAF=Global Assessment of Functioning. PANSS=Positive and Negative Syndrome Scale.

## Author contributions

Conceptualization: MK, LR, DK

Methodology: MK, LR, DK, KS, SP, PF, AZ, MZ

Investigation: MK, LR, DK, KS, SP

Visualization: MK, LR, DK, FR

Funding acquisition: PF, AS, DK, FR, EW

Project administration: DK, FR, EW, AS, PF

Data Collection: MK, AH, MSK, VM, EB, VY, JM

Data Processing: MK, LR, DK, SP, TJ, BK, GH, MSK, VM, AH

Supervision: MK, LR, DK

Writing – original draft: MK, LR, DK

Writing – review & editing: MK, LR, DK, KS, AZ, MZ, FR, EB

## CDP Working Group

Stephanie Behrens, Emanuel Boudriot, Man-Hsin Chang, Valéria de Almeida, Sylvia de Jonge, Fanny Dengl, Peter Falkai, Laura E. Fischer, Nadja Gabellini, Vanessa Gabriel, Sabrina Galinski, Thomas Geyer, Katharina Hanken, Alkomiet Hasan, Genc Hasanaj, Alexandra Hisch, Georgios Ioannou, Iris Jäger, Marcel Kallweit, Temmuz Karali, Susanne Karch, Berkhan Karslı, Daniel Keeser, Christoph Kern, Nicole L. Klimas, Maxim Korman, Nikolaos Koutsouleris, Lenka Krcmar, Verena Meisinger, Julian Melcher, Matin Mortazavi, Joanna Moussiopoulou, Karin Neumeier, Frank Padberg, Boris Papazov, Irina Papazova, Sergi Papiol, Pauline Pingen, Oliver Pogarell, Siegfried G. Priglinger, Florian J. Raabe, Lukas Roell, Moritz J. Rossner, Philipp Sämann, Zhuanghua Shi, Andrea Schmitt, Susanne Schmölz, Eva C. Schulte, Enrico Schulz, Benedikt Schworm, Elias Wagner, Sven Wichert, Vladislav Yakimov, Peter Zill

## Funding

The procurement of the MRI scanner was supported by the Deutsche Forschungsgemeinschaft (DFG, German Research Foundation) grant for major research (DFG, INST 86/1739-1 FUGG). This research was supported by BMBF with the EraNet project GDNF UpReg (01EW2206) to PF, AS, VY and GH. The study is funded by the EU HORIZON-INFRA-2024-TECH-01-04 project DTRIP4H 101188432 to PF, AS and FR. VY was supported by the Residency/PhD track of the International Max Planck Research School for Translational Psychiatry (IMPRS-TP). VY is supported by the Faculty of Medicine at LMU Munich (FöFoLe Reg.-Nr. 1226/2024). JM was supported by the Faculty of Medicine at LMU Munich (FöFoLe Reg.-Nr. 1167). The study was endorsed by the Federal Ministry of Education and Research (Bundesministerium für Bildung und Forschung [BMBF]) within the initial phase of the German Center for Mental Health (DZPG) (grant: 01EE2303A, 01EE2303F to PF). The study was funded by the Supplement to BMBF funding for the German Centre for Mental Health (DZPG) by the Bavarian State Ministry for Science and the Arts with the Grant for the research project ‘Improving Infrastructures for DZPG and NAKO Cohorts” to PF, DK and BK.

## Competing interests

PF received speaker fees from Boehringer Ingelheim, Janssen, Otsuka, Lundbeck, Recordati, and Richter and was a member of the advisory boards of these companies and Rovi. EW was invited to advisory boards from Recordati, Teva and Boehringer Ingelheim. MZ received speaker fees from Novartis Pharma GmbH. All other authors report no potential conflicts of interest.

## Data and materials availability

Clinical scores, neuropsychological and structural T1w and T2w MRI data were shared with the Open Science initiative Psy-ShareD and are publicly available there (https://psyshared.com/). The resting state and DTI data are being prepared for sharing with ENIGMA.

## Acknowledgments

Data collection and sharing for this project was provided by the Human Connectome Project (HCP; Principal Investigators: Bruce Rosen, M.D., Ph.D., Arthur W. Toga, Ph.D., Van J. Weeden, MD). HCP funding was provided by the National Institute of Dental and Craniofacial Research (NIDCR), the National Institute of Mental Health (NIMH), and the National Institute of Neurological Disorders and Stroke (NINDS). HCP data are disseminated by the Laboratory of Neuroimaging at the University of Southern California. Access to the HCP Ageing data was given to DK and LR through an application via the MIMH Data Archive.

Large Language Models (GPT-4 and Claude 3.7) were used to support coding efforts (prompt: "We are trying to calculate X with code Y and receive error Z. Can you comment on the error and suggest a solution? Elaborate on your solution.") and polish the writing (prompt: "This paragraph feels a bit clunky, can you suggest an improved version appropriate for scientific writing?”).

## Notes

### Clinical Protocols

https://www.frontiersin.org/journals/psychiatry/articles/10.3389/fpsyt.2023.1179811/full

### Author Declarations

This project was conducted as part of the Clinical Deep Phenotyping study, an extension of the Mental Health Biobank (ethics project No. 18 716), which received approval from the ethics committee of the Faculty of Medicine, Ludwig Maximilian University Munich (project Nos. 20 0528 and 22 0035) and registered at the German Clinical Trials Register (DRKS00024177)

